# Vaccination against SARS-CoV-2 in UK school-aged children and young people decreases infection rates and reduces COVID-19 symptoms

**DOI:** 10.1101/2022.03.13.22272176

**Authors:** Erika Molteni, Liane S. Canas, Kerstin Kläser, Jie Deng, Sunil S. Bhopal, Robert C. Hughes, Liyuan Chen, Benjamin Murray, Eric Kerfoot, Michela Antonelli, Carole H. Sudre, Joan Capdevila Pujol, Lorenzo Polidori, Anna May, Alexander Hammers, Jonathan Wolf, Tim D. Spector, Claire J. Steves, Sebastien Ourselin, Michael Absoud, Marc Modat, Emma L. Duncan

## Abstract

**Background:** We aimed to explore the effectiveness of one-dose BNT162b2 vaccination upon SARS-CoV-2 infection, its effect on COVID-19 presentation, and post-vaccination symptoms in children and young people (CYP) in the UK during periods of Delta and Omicron variant predominance.

**Methods:** In this prospective longitudinal cohort study, we analysed data from 115,775 CYP aged 12-17 years, proxy-reported through the Covid Symptom Study (CSS) smartphone application. We calculated post-vaccination infection risk after one dose of BNT162b2, and described the illness profile of CYP with post-vaccination SARS- CoV-2 infection, compared to unvaccinated CYP, and post-vaccination side-effects.

**Findings:** Between August 5, 2021 and February 14, 2022, 25,971 UK CYP aged 12-17 years received one dose of BNT162b2 vaccine. Vaccination reduced (proxy-reported) infection risk (-80·4% and -53·7% at 14–30 days with Delta and Omicron variants respectively, and -61·5% and -63·7% after 61–90 days). The probability of remaining infection-free diverged soon after vaccination, and was greater in CYP with prior SARS-CoV-2 infection. Vaccinated CYP who contracted SARS-CoV-2 during the Delta period had milder disease than unvaccinated CYP; during the Omicron period this was only evident in children aged 12-15 years. Overall disease profile was similar in both vaccinated and unvaccinated CYP. Post-vaccination local side-effects were common, systemic side-effects were uncommon, and both resolved quickly.

**Interpretation:** One dose of BNT162b2 vaccine reduced risk of SARS-CoV-2 infection for at least 90 days in CYP aged 12-17 years. Vaccine protection varied for SARS-CoV-2 variant type (lower for Omicron than Delta variant), and was enhanced by pre-vaccination SARS-CoV-2 infection. Severity of COVID-19 presentation after vaccination was generally milder, although unvaccinated CYP also had generally mild disease. Overall, vaccination was well-tolerated.

**Funding:** UK Government Department of Health and Social Care, Chronic Disease Research Foundation, The Wellcome Trust, UK Engineering and Physical Sciences Research Council, UK Research and Innovation London Medical Imaging & Artificial Intelligence Centre for Value Based Healthcare, UK National Institute for Health Research, UK Medical Research Council, British Heart Foundation and Alzheimer’s Society, and ZOE Limited.

**Research in context:** 

**Evidence before this study:** We searched PubMed database for peer-reviewed articles and medRxiv for preprint papers, published between January 1, 2021 and February 15, 2022 using keywords (“SARS-CoV-2” OR “COVID-19”) AND (child* OR p?ediatric* OR teenager*) AND (“vaccin*” OR “immunization campaign”) AND (“efficacy” OR “effectiveness” OR “symptoms”) AND (“delta” or “omicron” OR “B.1.617.2” OR “B.1.1.529”). The PubMed search retrieved 36 studies, of which fewer than 30% specifically investigated individuals <18 years.

Eleven studies explored SARS-CoV-2 viral transmission: seroprevalence in children (n=4), including age-dependency of susceptibility to SARS-CoV-2 infection (n=1), SARS-CoV-2 transmission in schools (n=5), and the effect of school closure on viral transmission (n=1).

Eighteen documents reported clinical aspects, including manifestation of infection (n=13), symptomatology, disease duration, and severity in children. Other studies estimated emergency department visits, hospitalization, need for intensive care, and/or deaths in children (n=4), and explored prognostic factors (n=1).

Thirteen studies explored vaccination-related aspects, including vaccination of children within specific paediatric co-morbidity groups (e.g., children with Down syndrome, inflammatory bowel disease, and cancer survivors, n=4), mRNA vaccine efficacy in children and adolescents from the general population (n=7), and the relation between vaccination and severity of disease and hospitalization cases (n=2). Four clinical trials were conducted using mRNA vaccines in minors, also exploring side effects. Sixty percent of children were found to have side effects after BNT162b2 vaccination, and especially after the second dose; however, most symptoms were mild and transient apart from rare uncomplicated skin ulcers. Two studies focused on severe adverse effects and safety of SARS-CoV-2 vaccines in children, reporting on myocarditis episodes and two cases of Guillain-Barrè syndrome. All other studies were beyond the scope of our research.

**Added value of this study:** We assessed multiple components of the UK vaccination campaign in a cohort of children and young people (CYP) aged 12-17 years drawn from a large UK community-based citizen-science study, who received a first dose of BNT162b2 vaccine. We describe a variant-dependent protective effect of the first dose against both Delta and Omicron, with additional protective effect of pre-vaccination SARS-CoV-2 infection on post-vaccination re-infection. We compare the illness profile in CYP infected post-vaccination with that of unvaccinated CYP, demonstrating overall milder disease with fewer symptoms for vaccinated CYP. We describe local and systemic side-effects during the first week following first-dose vaccination, confirming that local symptoms are common, systemic symptoms uncommon, and both usually transient.

**Implications of all the available evidence:** Our data confirm that first dose BNT162b2 vaccination in CYP reduces risk of infection by SARS-CoV-2 variants, with generally local and brief side-effects. If infected after vaccination, COVID-19 is milder, if manifest at all. The study aims to contribute quantitative evidence to the risk-benefit evaluation of vaccination in CYP to inform discussion regarding rationale for their vaccination and the designing of national immunisation campaigns for this age group; and applies citizen-science approaches in the conduct of epidemiological surveillance and data collection in the UK community.

Importantly, this study was conducted during Delta and Omicron predominance in UK; specificity of vaccine efficacy to variants is also illustrated; and results may not be generalizable to future SARS-CoV-2 strains.

## Introduction

SARS-CoV-2 infection in children and young people (CYP) is asymptomatic in approximately 50% of cases ^1, 2^ ; and in symptomatic cases is usually mild, of short duration, with low risk of hospitalisation and/or death ^3, 4^. In the general population, mass vaccination against SARS-CoV-2 is a key strategy to reduce morbidity and mortality of COVID-19, and manage demand on healthcare systems ^5^. Considering vaccination in children specifically, two-dose vaccination with BNT162b2 was highly efficacious in reducing infection risk in children aged 5-15 years, with most side-effects mild and short-lived, in studies conducted pre-Delta variant predominance ^6, 7^. However, concerns regarding vaccine-related risk of myo- and peri-carditis particularly in boys and young men have subsequently emerged ^8–10^.

SARS-CoV-2 vaccination campaigns in CYP have varied internationally. Evaluating vaccination for children includes consideration of the effect of vaccination upon susceptibility and severity of SARS-CoV-2 infection, of the strong correlation of age and COVID-19 severity, and of the educational and psychosocial effects of ongoing outbreaks, lock-downs, and quarantine policies (e.g., https://www.cdc.gov/coronavirus/2019-ncov/vaccines/recommendations/children-teens.html). In the UK, universal single-dose vaccination of CYP was authorized for the BNT162b2 vaccine for young people aged 16-17 years on August 4, 2021 (https://www.gov.uk/government/news/jcvi-issues-updated-advice-on-covid-19-vaccination-of-young-people-aged-16-to-17) and extended to children aged 12-15 years on September 13, 2021 (https://www.gov.uk/government/publications/universal-vaccination-of-children-and-young-people-aged-12-to-15-years-against-covid-19/universal-vaccination-of-children-and-young-people-aged-12-to-15-years-against-covid-19) with primary care advice received shortly thereafter. When these policies were announced, Delta (B.1.617.2) was the predominant circulating variant in the UK. After November 27, 2021, the Omicron (B.1.1.529) variant emerged and spread rapidly, becoming the predominant variant in the UK after December 20, 2021 (https://assets.publishing.service.gov.uk/government/uploads/system/uploads/attachment_data/file/1043807/technical-briefing-33.pdf).

Estimates of vaccine effectiveness in real-world settings may differ from trial data, with changes over time in exposure, prior infection-induced immunity, emergence of new variants, testing policies, and ‘track and trace’ strategies. Real-world studies in individuals aged 12-18 years confirmed high BNT162b2 vaccine effectiveness against severe COVID-19 presentation ^11^. However, data on vaccination effect upon community-managed infections, and on symptom presentation, are less clear for the school-aged population. Moreover, effectiveness of vaccination against newer SARS-CoV-2 variants remains incompletely determined ^12^.

Citizen science allows capturing of phase 4 data for medicinal products after their pivotal phase 3 trials, allowing real-time and real-world monitoring of effectiveness and side-effect profile, at scale ^13, 14^. Here, using such an approach, we aimed to explore effectiveness of first-dose BNT162b2 vaccination (i.e., the UK vaccination policy in CYP at the time of this study) upon risk of subsequent SARS-CoV-2 infection for individuals aged 12 to 17 years. We considered also the effects for two variants, Delta and Omicron, and of SARS-CoV-2 infection prior to first-dose vaccination. We also studied the effect of vaccination on COVID-19 presentation, including hospital presentation; and post-vaccination symptomatology in children and young people.

## Methods

This prospective observational study used data from the COVID Symptom Study, collected through a smart-phone application: the ZOE COVID Study App (details previously published ^14^). Briefly, adult participants self-report symptoms, including responses to direct questions (Supplementary Table 1) and free-text, any SARS-CoV- 2 testing and results, vaccination, post-vaccination symptoms (Supplementary Table 2), and health care access, as well as demographic and co-morbidity data (the latter mainly informed by common adult co-morbidities). Adult contributors can proxy-report for others, including children; however, data cannot be linked between contributor and proxy-reported individual. Teenagers aged 16-17 can also self-report.

Ethics approval was granted by the KCL Ethics Committee (LRS-19/20-18210). Upon registration with the app, all participants were provided with study information, and gave consent for their data to be used for COVID-19 research. Governance was specifically granted to enable use of proxy-reported data, which included data from children. Research was conducted in full compliance with the Declaration of Helsinki and further updates.

Data from all proxy-reported UK individuals aged 12-17 years (12-15 years: herein termed *children*; 16-17 years: herein termed *young people*) were considered from August 5, 2021 (i.e., the day after announcement of the UK universal vaccination policy of teenagers aged 16-17 years) to February 14, 2022 (census date), extracted through ExeTera software ^15^. Self-reported data were excluded, to avoid potential duplicate data entry (i.e., from self- and proxy-reporting of older teenagers) and because our previous analysis showed overall lower perseverance and reporting density by self-reporting teenagers compared with proxy-reported teenagers ^3^. Some vulnerable CYP and those living with vulnerable adults were eligible to access vaccination earlier (July 19, 2021) and/or receive two vaccine doses (https://www.gov.uk/government/news/jcvi-issues-advice-on-covid-19-vaccination-of-children-and-young-people). Data from these CYP were excluded by constraint upon vaccination date (i.e., after policy announcement dates, as above) and/or by vaccination dose number (i.e., only single/first dose data were considered). Thus, all analyses described herein refer to single first dose BNT162b2 vaccination, administered from August 5, 2021 to February 14, 2022.

Study design is depicted in Figure 1, with inclusion/exclusion criteria and pertinent numbers for each sub-analysis presented in Supplementary Table 3. Prior SARS-CoV-2 infection was defined as a previous proxy-logged positive test for SARS-CoV-2 (either reverse transcription polymerase chain reaction (PCR) or lateral flow antigen test (LFAT) or a declaration of previous infection at time of first registration in the app with or without confirmatory test information. Prior SARS-CoV-2 infection was defined relative to date of vaccination, or to date of matching for unvaccinated controls as per each analysis (described below). Testing for SARS-CoV-2 was per local guidelines, and may have been prompted by symptoms, school and/or other social requirements, contact tracing, or other reason; rationale for testing for proxy-reported individuals was not captured.

**Figure 1.**
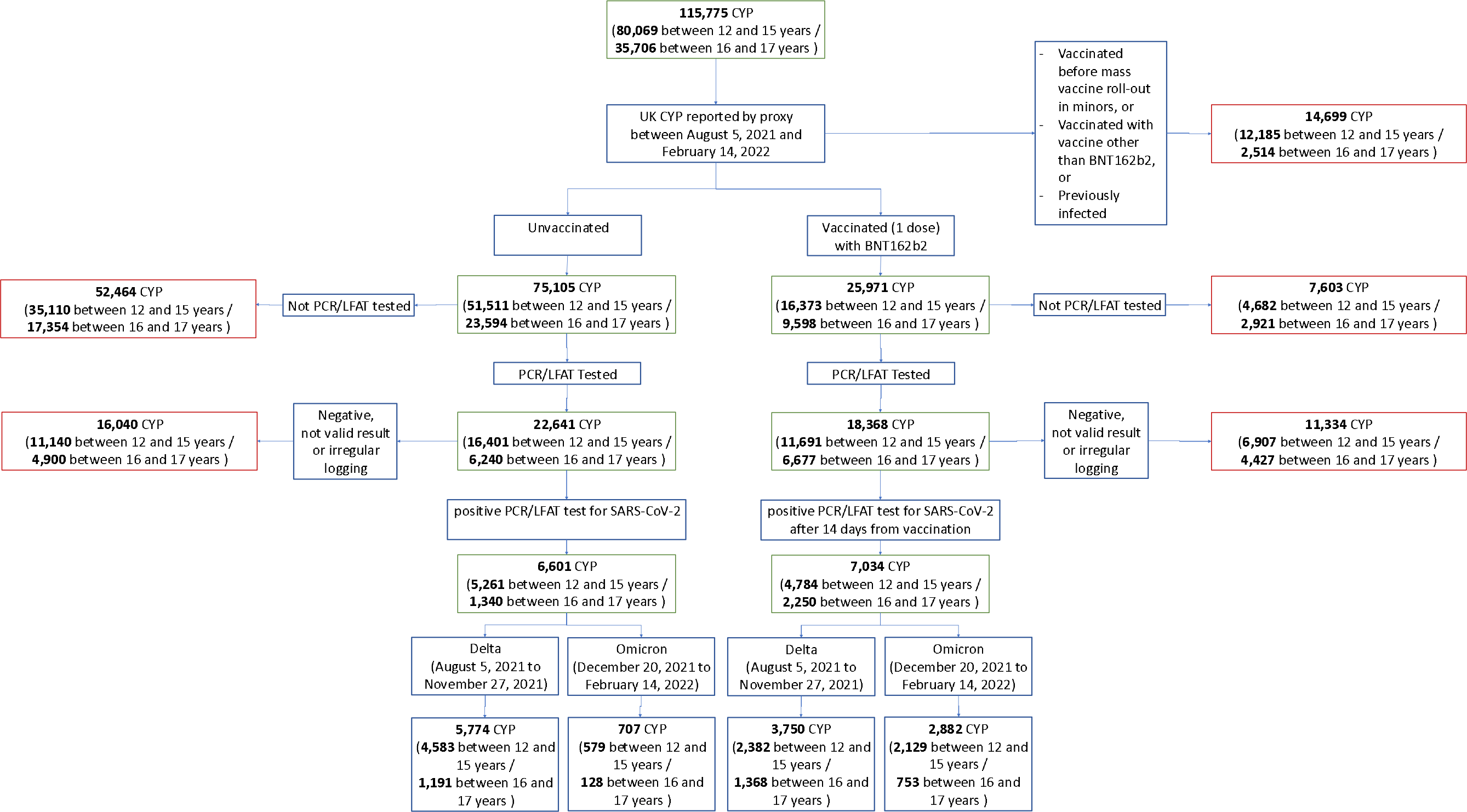
Flowchart of participants according to inclusion and exclusion criteria for this study. Entire cohort is given first, before subdivision into vaccinated and unvaccinated CYP. Note here the two age groups had differing time periods for data consideration (young people aged 16–17 years: able to receive vaccination after August 4, 2021; children aged 12-15 years: able to receive vaccination after September 18, 2021). Further subdivision provides numbers of individuals for periods of Delta and Omicron variant predominance in UK. Considered timeframes for Delta and Omicron variant predominance were not contiguous; thus, numbers of CYP within each timeframe do not sum to numbers across the entire study period. Not valid result = test result proxy-reported as “failed test” or “still waiting”. Irregular logging = proxy-reporting with intervals of more than 7 days between proxy-reports during illness. PCR – polymerase chain reaction. LFAT – lateral flow antigen test.

### Post-vaccination (proxy-reported) infection risk in CYP without prior SARS-CoV-2 infection

Data from all UK proxy-reported CYP aged 12-17 years tested for SARS-CoV-2 (PCR or LFAT) logged between August 5, 2021 and February 14, 2022 were considered.

Data were also considered within two time frames: August 5, 2021 to November 27, 2021, when Delta was the predominant circulating SARS-CoV-2 variant (minimum prevalence >70%); and December 20, 2021 to February 14, 2022, when Omicron was prevailing (minimum prevalence >70%) (https://www.gisaid.org/hcov19-variants/).

To assess real-world effectiveness, vaccinated and unvaccinated CYP naïve to SARS-CoV-2 were compared. To allow time for induction of immunity ^16^, test results for vaccinated CYP were considered from 14 days after vaccination (as previously^17^). Unvaccinated CYP were excluded if they had tested positive before 4 August and were only considered until the first positive test result was recorded (if such existed).

For each CYP (vaccinated or unvaccinated), all negative tests until the first positive test (included, if such existed) were considered; after the first positive test, all subsequent tests were ignored. As previously ^17^, test results were grouped by week: for CYP with at least one negative and one positive test within a single week, the positive test was used. In case of multiple negative tests within one week, the last one was used.

To control for viral prevalence and strain, vaccinated CYP were matched with the unvaccinated by week of testing. As previously ^17^, an unvaccinated CYP could serve as a control for more than one week, if tested within each corresponding week.

We used Poisson regression to compare weekly incidence of SARS-CoV-2 infection in the vaccinated and unvaccinated CYP, adjusting for number of tests, age, number of co-morbidities, sex, and weekly incidence of infection nationally (by controlling for week of testing through a categorical variable). We obtained the adjusted risk reduction as RR=(riskratio_n_−1)*100, where n encodes the week considered and *riskratio* is the ratio of infection rates in vaccinated vs. unvaccinated individuals, estimated through the Poisson model. Descriptive statistics are also presented.

### Risk (hazard) of post-vaccination infection in vaccinated and unvaccinated CYP without prior SARS-CoV-2 infection

To assess effect of vaccination upon remaining infection-free (*survival*) in SARS-CoV- 2-naïve individuals, we matched 1:1 vaccinated and unvaccinated CYP for week of test, sex, and age, excluding CYP with prior SARS-CoV-2 infection. For this analysis, *no prior infection* was defined in vaccinated CYP as no infection before vaccination; and for unvaccinated CYP as no prior infection before week of matching. We constructed Kaplan-Meier survival curves to SARS-CoV-2 tests, with the first positive test being the failure event. Analysis considered periods of Delta and Omicron variant predominance.

### Effect of pre-vaccination SARS-CoV-2 infection on post-vaccination infection risk

To assess the effect of prior SARS-CoV-2 infection we matched 1:1 vaccinated CYP with vs. without prior SARS-CoV-2 infection for week of test, sex, and age. Kaplan-Meier survival curves were constructed. Analysis considered periods of Delta and Omicron variant predominance, as defined above.

### Illness profile in vaccinated vs. unvaccinated CYP with SARS-CoV-2: CYP without prior infection

We assessed symptom manifestation and illness burden in SARS-CoV-2-naïve vaccinated CYP, who subsequently tested positive, including symptomatic and asymptomatic CYP. Symptoms were considered from one week before to one month after a positive test, with the additional constraint that symptoms had to commence between one week before and two weeks after the positive test result (as previously^3^). We defined disease burden as the median count of individual symptoms per week.

We calculated illness duration from the first day of first symptom to the last day of last symptom, and assessed prevalence and distribution of duration for each symptom (as previously ^3^).

We then compared illness profile in vaccinated vs. unvaccinated CYP without prior infection, matched for week of test, sex and age ^3^. Matching was performed by Euclidean distance, with equal number of subjects in each population when the number of unvaccinated subjects was sufficient; however, this was not possible for all comparisons due to rapidly declining numbers of unvaccinated CYP over time. Here prior infection was defined in vaccinated CYP as no prior infection before date of vaccination and in unvaccinated CYP as no prior infection by week of matching. Asymptomatic subjects who tested positive were included in this analysis. As previously ^18^, after adjustment for sex, age, and number of comorbidities, odds ratios (ORs) were computed for each symptom between the two groups, with significance adjusted for False Discovery Rate. Hospital presentation (emergency room presentation and/or hospital admission) was also assessed, noting here that we did not have health records linkage and thus could not determine reason for presentation.

### Post-vaccination symptoms

We assessed post-vaccination symptoms in all vaccinated CYP. For this analysis only, we excluded CYP with a positive SARS-CoV-2 test within three months prior to vaccination, so that reported symptoms pertained to vaccination *per se* rather than residual COVID-19. As previously ^17^, we counted the number of individuals experiencing any symptom in the seven days after vaccination, assessing daily prevalence of pre-specified systemic (Supplementary Table 1) and local (Supplementary Table 2) symptoms during this time period.

A technical issue in app software occurred on August 5, 2021 which prevented recording of local symptoms, which was resolved by August 23, 2021. Thus CYP due to report post-vaccination local symptoms within this time window (n=357) were removed from the analysis of local symptoms; systemic symptoms were still able to be assessed throughout. Notwithstanding this technical issue, reporting of systemic and/or local symptoms could occur independently; thus, the numbers of CYP assessed for systemic and local symptoms differ even outside the August 5-23 window, as reported in Table 3 and Supplementary Table 3.

Lastly, free text field entries were analysed for symptoms not queried through the standard questions. Severe side-effects of BNT162b2 vaccine reported in the literature were specifically assessed within the free text, including anaphylaxis, thrombosis, cytopenia, myocarditis, pericarditis, Guillain-Barré syndrome, and (multi)organ failure ^8–10, 19^.

## Results

### Cohort Description

The overall cohort comprised 25,971 vaccinated and 75,105 unvaccinated CYP: 9,598 vaccinated and 23,594 unvaccinated young people (aged 16-17 years) proxy-reported between August 5, 2021 and February 14, 2022; and 16,373 vaccinated and 51,511 unvaccinated children (aged 12-15 years) between September 18, 2021 and February 14, 2022 (Figure 1, Table 1).

**Table 1.** Demographic characteristics of UK CYP proxy-logged as part of the COVID Symptom Study between August 5, 2021 and February 14, 2022.

|  | **Vaccinated CYP**<br>**(first dose BNT162b2)** |  | **Unvaccinated CYP** |  |
| --- | --- | --- | --- | --- |
|  | ***Children (aged***<br>***12-15 years)*** | ***Young people***<br>***(aged 16-17***<br>***years)*** | ***Children (aged***<br>***12-15 years)*** | ***Young people***<br>***(aged 16-17***<br>***years)*** |
| **Number of CYP proxy-reported within relevant time frames\*** | 16,373 | 9,598 | 51,511 | 23,594 |
| **Number of CYP tested for SARS-CoV-2 (n)** | 11,691 | 6,677 | 16,401 | 6,240 |
| **#Positive test result (n (%))** | 4,784 (40.9) | 2,250 (33.7) | 5,261 (32.1) | 1,340 (21.5) |
| **#Age (mean (SD))** | 13.58 (1.09) | 16.46 (0.50) | 13.54 (1.11) | 16.43 (0.50) |
| **#BMI (mean (SD))** | 19.66 (3.67) | 20.80 (3.86) | 19.75 (3.76) | 20.72 (3.82) |
| **#Male (n (%))** | 6,105 (52.2) | 3,508 (52.5) | 8,615 (52.5) | 3,310 (53.0) |
| **#CYP with at least one comorbidity (n (%))** | 644 (5.5) | 362 (5.4) | 917 (5.6) | 340 (5.4) |

\*Time frames: For young people aged 16-17 years, data were considered between August 5, 2021 and February 14, 2022. For children aged 12-15 years, data were considered between September 18, 2021 and February 14, 2022.

Data from 6,677 vaccinated young people with at least one post-vaccination PCR or LFAT test were compared to 6,240 unvaccinated young people analogously proxy-reported between August 5, 2021 and February 14, 2022. Similarly, data from 11,691 vaccinated children with at least one test post-vaccination were compared to 16,401 unvaccinated children, between September 18, 2021 and February 14, 2022.

Significantly fewer unvaccinated CYP were tested, compared with vaccinated CYP (Table 1: assessed by Chi-squared testing; p<0.00001 for both young people and children). Other demographics are presented in Table 1. Cohort descriptions and sample size for each analysis are detailed in Supplementary Table 3.

### Post-vaccination infection risk

During the period of Delta variant predominance, and for SARS-CoV-2-naïve CYP, the adjusted risk ratio of infection post-vaccination vs no vaccination was -80·4% (95%CI: -82·2 to -78·5) 14-30 days post-vaccination; -86·4% (95%CI: -88·7 to -83·5) one to two months post-vaccination, and -61·5% (95%CI: -73·7 to -43·5) two to three months post-vaccination (Figure 2: left panel). During the Omicron period, the adjusted risk ratios were -53·7% (95%CI: -62·2 to -43·3), -57·9% (95%CI: -63·9 to - 50·9), and -63·7% (95%CI: -67·9 to -59·0), respectively. (Figure 2: right panel).

**Figure 2.**
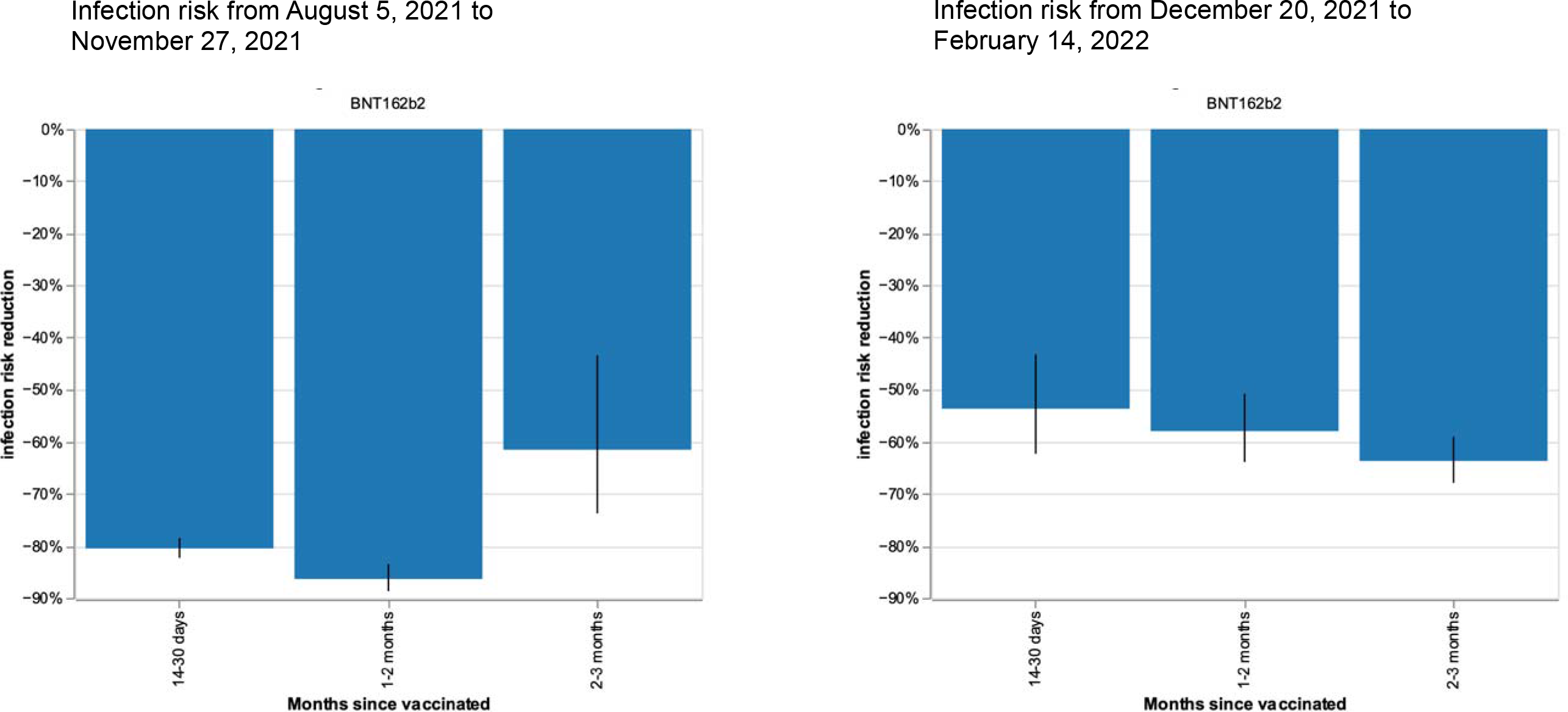
Infection risk reduction after single vaccination with BNT162b2 in 12- to17-year-old CYP during periods of Delta (left) and Omicron (right) variant predominance in UK. Bars represent risk reduction at 14-30 days, 1-2 months, and 2-3 months for post-vaccination infection, compared with unvaccinated CYP. The black lines show 95% CIs. Number of observations (i.e., tests) of CYP aged 12-17 years: for period of Delta variant predominance n (test) =15,308 (4,793 unvaccinated, 10,515 vaccinated); for period of Omicron variant predominance n (test)=8,203 (930 unvaccinated, 7,273 vaccinated).

Breakdowns by age groups for Delta and Omicron periods are shown in Supplementary Figures 1 and 2 respectively. Comparing periods of Delta and Omicron predominance, risk reduction after vaccination appeared higher during Delta compared to Omicron, at least for the first three months in young people aged 16-17 years (Supplementary Figure 1), and at least for the first two months after vaccination in children aged 12-15 years (Supplementary Figure 2).

### Risk (hazard) of post-vaccination infection over time

A protective effect of vaccination on SARS-CoV-2 infection was evident promptly, with risk quickly diverging between vaccinated and unvaccinated CYP as shown in the Kaplan-Meier survival analysis (Figure 3).

**Figure 3.**
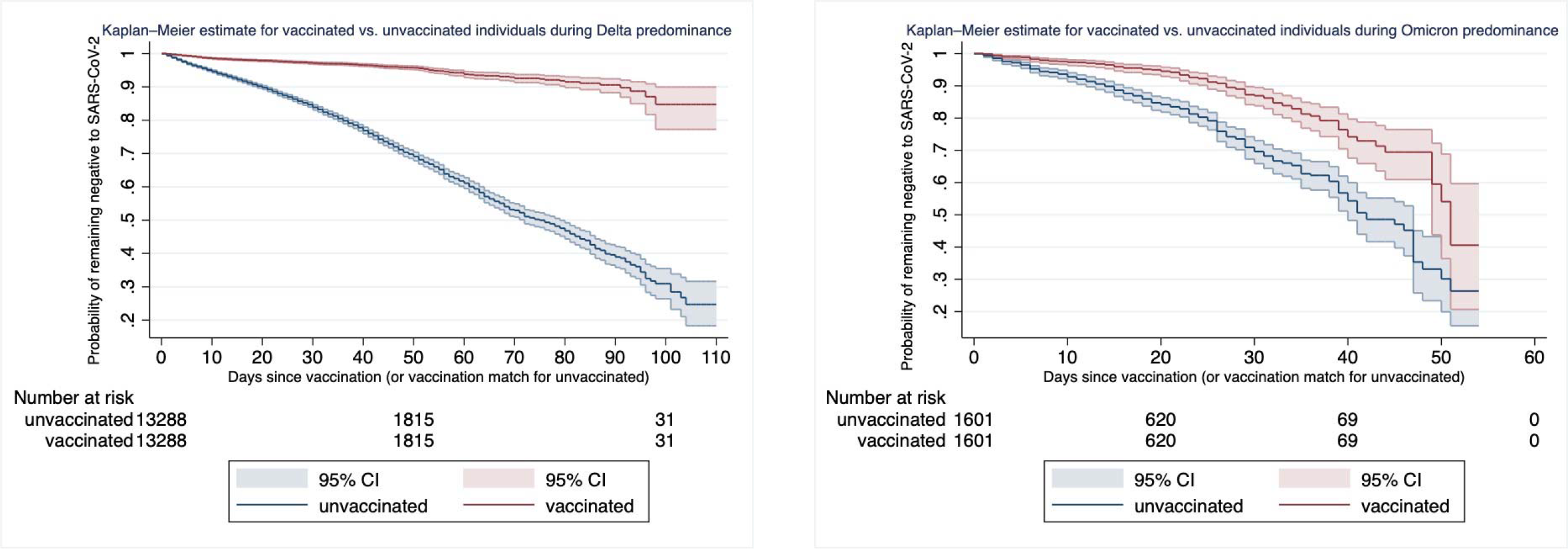
Survival analysis: Kaplan-Meier plots showing probability of remaining free from SARS-CoV-2 infection over time for

In CYP without prior SARS-CoV-2 infection during Delta predominance, the probability of remaining infection-free post-vaccination was high: >90% for 11 weeks post-vaccination, decreasing just below 90% at 90 days (Figure 3: left panel). During Omicron predominance, the probability of remaining infection-free post-vaccination was shorter: >90% until 25 days post-vaccination, <90% at 30 days, and <80% at 40 days. Additionally, the vaccine-induced risk reduction was smaller for vaccinated vs. unvaccinated CYP during Omicron vs. Delta periods.

Considering vaccinated CYP with vs. without prior SARS-CoV-2 infection: prior infection enhanced the protective effect of vaccination against infection. During Delta predominance, the risk of reinfection was close to null beyond 100 days (Figure 4, left panel). During Omicron predominance, prior infection continued to confer enhanced vaccine efficacy; however, data were insufficient to provide a robust quantitative description (Figure 4, right panel).

**Figure 4.**
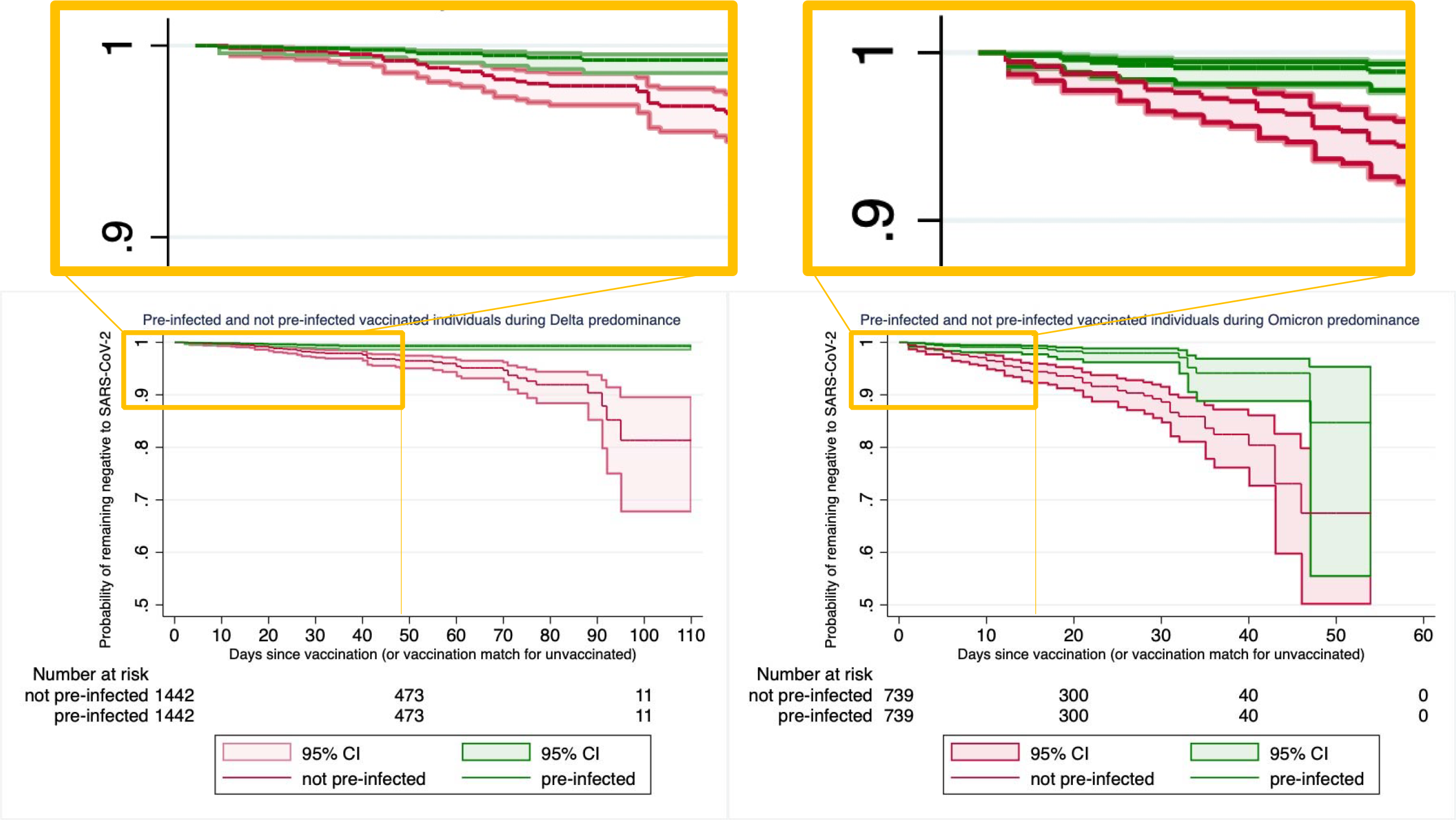
Survival analysis: Kaplan-Meier plots showing the probability of remaining free from SARS-CoV-2 infection over time for vaccinated CYP with or without SARS-CoV-2 infection prior to vaccination, during periods of Delta (left panel) and Omicron (right panel) variant predominance.

### Illness profile in CYP with *de novo* SARS-CoV-2 infection

During Delta predominance, all symptoms except rhinorrhoea and sneezing had lower prevalence in CYP infected post-vaccination (n=3,750) compared to infected unvaccinated CYP (n=5,774) (Figure 5, top panel), matched for age, gender, and BMI. Formal statistical testing showed lower odds ratios for multiple symptoms in vaccinated CYP including headache, fatigue, dizziness, eye soreness, arthralgias, low appetite, nausea; additionally, vaccinated children had lower odds for confusion and vaccinated young people had lower odds for multiple other symptoms including sore throat, persistent cough, fever, hoarse voice, myalgias, chest pain, abdominal pain, low mood, lymphadenopathy, diarrhoea, and rashes (Figure 6, left panel, Supplementary Table 4; alpha=0·05).

**Figure 5.**
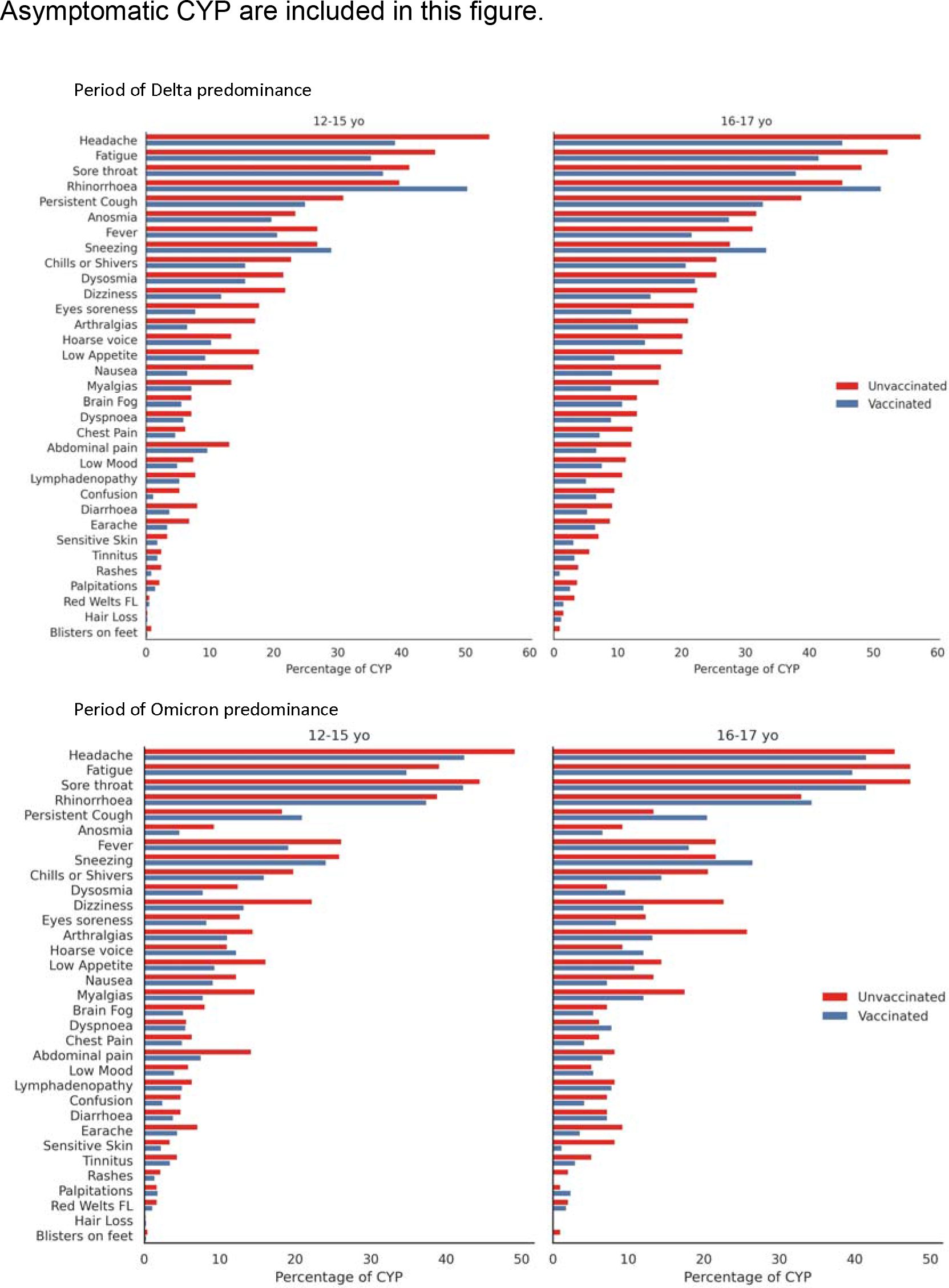
COVID-19 profile in vaccinated and unvaccinated CYP testing positive for SARS-CoV-2 during periods of Delta (top panels) and Omicron (bottom panels) variant predominance, in previously SARS-CoV-2-naïve children (left panels) and young people (right panels).

**Figure 6.**
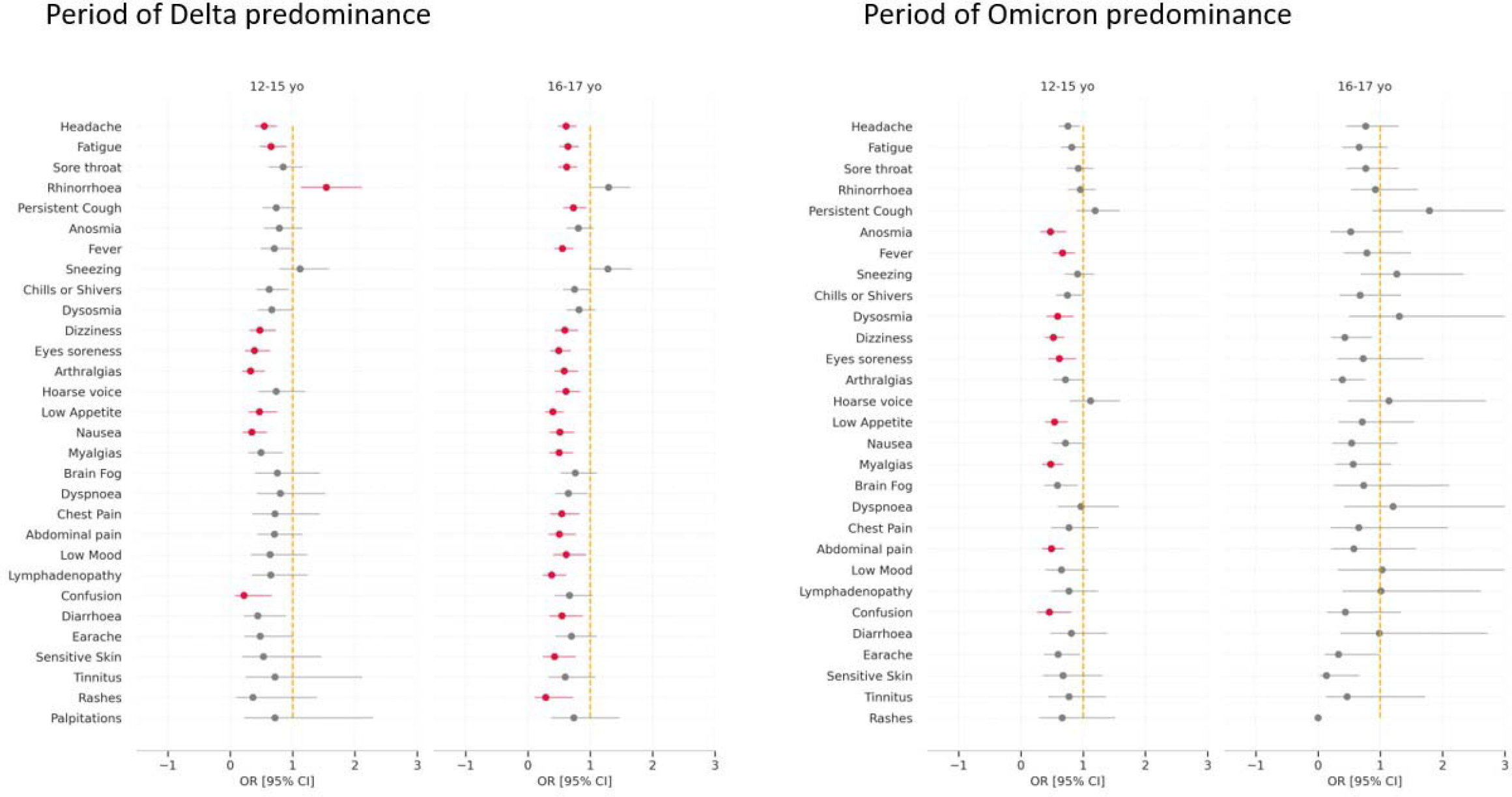
Odds ratios for symptom prevalence in vaccinated vs. unvaccinated CYP first testing positive for SARS-CoV-2 during periods of Delta (left) and Omicron (right) variant predominance. Vaccinated CYP had no reported SARS-CoV-2 infection prior to vaccination. Asymptomatic CYP testing positive for SARS-CoV-2 are included in the computation. Red encodes significant p-values after false discovery rate correction (alpha =0.05).

Similarly, during Omicron predominance, most symptoms had lower prevalence in vaccinated (n=2,882) compared to unvaccinated CYP (n=707), except persistent cough and hoarse voice, and, in young people only, rhinorrhoea, sneezing, dysosmia, dyspnoea, and low mood, noting the marginal difference in these latter infrequent symptoms (Figure 5, bottom panel). Formal testing showed lower odds ratios for anosmia, fever, dysosmia, dizziness, eye soreness, low appetite, myalgias, abdominal pain and confusion in children, but no significant differences for any symptom in young people (Figure 6, right panel, Supplementary Table 4).

For both Delta and Omicron periods, median symptom duration was 1 day for most symptoms, in both vaccinated and unvaccinated CYP, other than very rare cutaneous manifestations (welts, blisters, hair loss), and eye soreness in young people with Omicron infection (Supplementary Figure 3). Overall, symptom persistence was similar during Delta vs. Omicron predominance and for vaccinated and unvaccinated CYP (Supplementary Figures 4 and 5 for children and young people, respectively).

Burden of illness was assessed for the first week and the first 28 days. This analysis incorporated both symptomatic and asymptomatic test-positive individuals (Table 2: descriptive data only). During Delta predominance, vaccinated CYP had slightly fewer symptoms than unvaccinated CYP both the first week and the first 28 days; total illness duration was similar. During Omicron predominance, symptom burden appeared globally slightly less than during the Delta period, and duration comparable. Symptom burden and illness duration were similar for vaccinated and unvaccinated CYP.

**Table 2.** Illness burden and duration, and hospitalisation numbers, for vaccinated and unvaccinated CYP with symptomatic de novo SARS-CoV-2 infection, for periods of Delta and Omicron variant predominance, and for the entire study period. Vaccinated CYP were considered from at least 14 days post-vaccination. Median and [IQR] are reported for illness burden (symptom count) and duration (days). Numbers (percentage) are reported for hospital presentation; CYP with a positive SARS-CoV-2 test may have presented to hospital for reasons other than COVID-19. Time frames for Delta and Omicron periods were not contiguous; thus, the joint two time periods do not correspond to the entire time period.

|  |  | Delta Period (5<sup>th</sup> August – 27<sup>th</sup> November 2021) |  |  |  | Omicron Period (20<sup>th</sup> December 2021 – 14<sup>th</sup> February 2022) |  |  |  | Entire time period of study (5<sup>th</sup> August 2021 – 14<sup>th</sup> February 2022) |  |  |  |
| --- | --- | --- | --- | --- | --- | --- | --- | --- | --- | --- | --- | --- | --- |
|  |  | Symptoms during first 7 days of illness (count, [IQR]) | Symptoms during first 28 days of illness (count [IQR]) | Duration of illness ((median [IQR]) | CYP presenting to hospital* (n (%)) | Symptoms during first 7 days of illness (count, [IQR]) | Symptoms during First 28 days of illness (count [IQR]) | Duration of illness (median [IQR]) | CYP presenting to hospital* (n (%)) | Symptoms during first 7 days of illness (count, [IQR]) | Symptoms during first 28 days of illness (count [IQR]) | Duration of illness (median [IQR]) | CYP presenting to hospital* (n (%)) |
| CYP: 12-15 years | Vaccinated | 4 symptoms [2; 7] | 3 symptoms [2; 4] | 2 days [1; 5] | 1 child (0.31%) | 4 symptoms [1; 7] | 2 symptoms [2; 3] | 2 days [1; 5] | 9 children (0.71%) | 4 symptoms [1; 7] | 2.5 symptoms [2; 3.75] | 2 days [1; 5] | 13 children (0.64%) |
|  | Unvaccinated | 6 symptoms [2.25; 10] | 4.75 symptoms [3.25; 5.38] | 1 day [1; 5] | 9 children (2.81%) | 4 symptoms [1; 7] | 3 symptoms [2; 4] | 1 day [1; 4] | 4 children (0.98%) | 5 symptoms [2; 8] | 4 symptoms [3.25; 5] | 2 days [1; 6] | 32 children (1.58%) |
| CYP: 16-17 years | Vaccinated | 5 symptoms [2; 8] | 4.25 symptoms [3; 5] | 3 days [1; 7] | 15 young people (2.66%) | 4 symptoms [0; 7] | 3 symptoms [1; 4] | 1 day [1; 4] | 1 young person (0.60%) | 4 s symptoms [1.5; 8] | 3.5 symptoms [3; 4] | 2 days [1; 7] | 17 young people (2.13%) |
|  | Unvaccinated | 7 symptoms [4; 11] | 5 symptoms [4.25; 6.75] | 3 days [1; 8] | 9 young people (1.60%) | 5 symptoms [2.5; 10] | 3 symptoms [0; 5] | 1 day [1; 3] | 1 young person (1.03%) | 7 symptoms [3; 11] | 5 symptoms [4.25; 6.38] | 3 days [1; 7] | 12 young people (1.50%) |

Very few CYP presented to the hospital during the entire study period. During the Delta period, 16 (1·8%) of 884 vaccinated CYP presented to hospital, compared to 18 (2·1%) of 864 unvaccinated. During the Omicron period, 10 (0·7%) of 1,435 vaccinated CYP presented to hospital, compared to 5 (1·0%) of 506 unvaccinated (Table 2). No statistical tests are possible for such low frequency data; and we are unable to determine reason for presentation (i.e., SARS-CoV-2 diagnosis may have been incidental).

### Post-vaccination symptoms in CYP without recent SARS-CoV-2 infection

Post-vaccination symptom prevalence are shown in Table 3.

**Table 3.** Vaccination side-effects reported at least once during the 7 days after vaccination, in UK CYP. Subjects positive to SARS-CoV-2 test prior to vaccination were excluded.

| **GENERAL POPULATION CHARACTERISTICS** |  |  |
| --- | --- | --- |
|  | **12-15-year-old group**<br>**(children)** | **16-17-year-old group**<br>**(young people)** |
| **SYSTEMIC SYMPTOMS** |  |  |
| Number of individuals considered for analysis of systemic symptoms (n) | 7,809 | 3,189 |
| One or more systemic symptom (n (%)) | 1,104 (14.1) | 266 (8.3) |
| Headache (n (%)) | 291 (3.7) | 63 (2.0) |
| Fatigue (n (%)) | 257 (3.3) | 57 (1.8) |
| Sore throat (n (%)) | 272 (3.5) | 54 (1.7) |
| Rhinorrhoea (n (%)) | 294 (3.8) | 48 (1.5) |
| Persistent cough (n (%)) | 116 (1.5) | 30 (0.9) |
| Anosmia/Dysosmia (n (%)) | 110 (1.4) | 30 (0.9) |
| Sneezing (n (%)) | 169 (2.2) | 28 (0.9) |
| Chills & Shivers (n (%)) | 104 (1.3) | 25 (0.8) |
| Nausea (n (%)) | 80 (1.0) | 21 (0.7) |
| Dizziness (n (%)) | 100 (1.3) | 20 (0.6) |
| Fever (n (%)) | 121 (1.5) | 19 (0.6) |
| Abdominal pain (n (%)) | 73 (0.9) | 15 (0.5) |
| Hoarse voice (n (%)) | 58 (0.7) | 15 (0.5) |
| Lymphadenopathy (n (%)) | 30 (0.4) | 14 (0.4) |
| Myalgia (n (%)) | 57 (0.7) | 11 (0.3) |
| Arthralgia (n (%)) | 62 (0.8) | 9 (0.3) |
| Low appetite (anorexia) (n (%)) | 52 (0.7) | 8 (0.3) |
| Dyspnoea (n (%)) | 27 (0.3) | 8 (0.3) |
| Earache (n (%)) | 26 (0.3) | 8 (0.3) |
| Low mood (depression) (n (%)) | 25 (0.3) | 7 (0.2) |
| Diarrhoea (n (%)) | 29 (0.4) | 6 (0.2) |
| Brain Fog (n (%)) | 31 (0.4) | 6 (0.2) |
| Delirium (n (%)) | 30 (0.4) | 6 (0.2) |
| Chest pain (n (%)) | 27 (0.3) | 4 (0.1) |
| Palpitations (n (%)) | 8 (0.1) | 4 (0.1) |
| Hair loss (n (%)) | 1 (<0.1) | 4 (0.1) |
| Tinnitus (n (%)) | 18 (0.2) | 3 (0.1) |
| **CUTANEOUS SYMPTOMS** |  |  |
| Eye Soreness | 35 (0.4) | 8 (0.3) |
| Skin Burning (n (%)) | 14 (0.2) | 3 (0.1) |
| Rash (n (%)) | 5 (0.1) | 2 (0.1) |
| Red welts on lips (n (%)) | 8 (0.1) | 1 (<0.1) |
| Blisters on feet | 2 (<0.1) | 0 (0.0) |
| **LOCAL SYMPTOMS** |  |  |
| Number of individuals considered for analysis of local symptoms | 7,751 | 2,876 |
| One or more local symptom (n (%)) | 5,485 (70.8) | 1,731 (60.2) |
| Tenderness (n (%)) | 3,557 (45.9) | 1,183 (41.1) |
| Pain (n (%)) | 3,240 (41.8) | 954 (33.2) |
| Warmth (n (%)) | 378 (4.9) | 128 (4.5) |
| Swelling (n (%)) | 388 (5.0) | 90 (3.1) |
| Redness (n (%)) | 298 (3.8) | 71 (2.5) |
| Axillary Lymphadenopathy (n (%)) | 114 (1.5) | 20 (0.7) |
| Pruritis (n (%)) | 88 (1.1) | 20 (0.7) |
| **Number of individuals** | 6 of 7,809 (0.1) | 0 of 3,189 (0.0) |
| **presenting to hospital post-**<br>**vaccination\* (n (%))** |  |  |

BMI is body mass index; SD is standard deviation.
\*Presentation to hospital: presenting to the emergency department and/or admission to hospital. Reason for presentation unknown.

Among children, 5,485 (70·8%) of 7,751 had one or more local symptom post-vaccination, most commonly tenderness (3,557 (45·9%) of 7,751 children) and local pain at the injection site (3,240 (41·8%)). 1,104 (14·1%) of 7,809 children had one or more systemic symptom, most commonly rhinorrhoea (294 (3·8%) of 7,809 children), headache (291 (3·7%), sore throat (272 (3·5%)), and fatigue (257 (3·3%)). Cutaneous symptoms were rare (<0·5%). Post-vaccination symptoms (both local and systemic) usually occurred on the first day post-vaccination, and resolved quickly in most children (Figure 7, upper panel).

**Figure 7.**
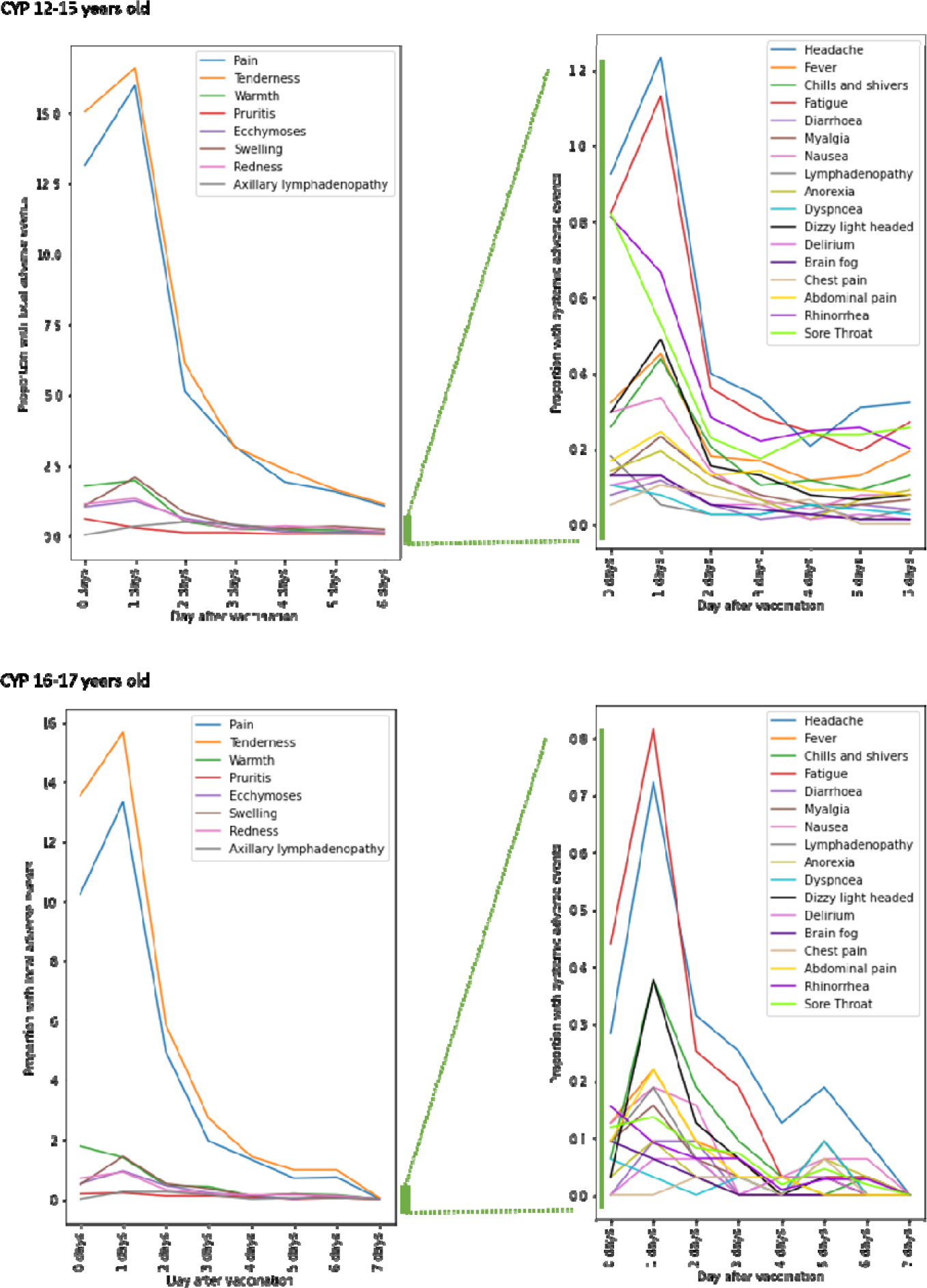
Proportion of CYP aged 12 to 17 years old, separated by age group, reporting vaccine side-effects after one dose of BNT162b2. Left panels: local symptoms at the arm of injection within 7 days; right panels: systemic symptoms presenting within the same 7 days. Data for children aged 12- 15 years are shown in upper panels and for young people aged 16-17 years in lower panels.

Among young people, 1,731 (60·2%) of 2,876 had one or more local symptom post-vaccination, again most commonly tenderness (1,183 (41·1%) of 2,876 young people) and local pain (954 (33·2%)). 266 (8·3%) of 3,189 had one or more systemic symptom, most commonly headache (63 (2·0%) of 3,189 young people), fatigue (57 (1·8%)), and sore throat (54 (1·7%)). Cutaneous symptoms were again rare (<0·4%). Similarly, both systemic and local symptoms usually occurred on the first day post-vaccination and resolved quickly (Figure 7, lower panel).

Supplementary Figure 6 shows post-vaccination local and systemic symptom profiles for the cohort overall.

Six (0·1%) of 7,809 children and none of the 3,189 young people presented to hospital after vaccination.

No question was asked specifically regarding myocarditis or pericarditis. Chest pain was reported in 27 (0.3%) of 7,809 children, and 4 (0.1%) of 3,189 young people; and dyspnoea in 27 (0.3%) of 7,809 children and 8 (0.3%) of 3,189 young people. Free-text scrutiny revealed five cases of tingling of the arm, hand, or fingers at the side of injection; two cases of insomnia; and one of lethargy. No reports were found of anaphylaxis, thrombosis, cytopenia, myocarditis, pericarditis, Guillain-Barré syndrome, or multi-organ failure. Analogously, we found no terms suggesting medical tests related to myocarditis or pericarditis, such as electrocardiography, echography, or creatinine kinase tests.

## Discussion

Here we show that vaccination with a single dose of BNT162b2 reduced risk of SARS-CoV-2 infection in CYP aged 12-17 years. The protective effect of vaccination was evident early, with risk of infection reduced 14 days post-vaccination and prompt divergence of Kaplan-Meier curves for infection-free survival. However, vaccine effectiveness may vary according to prevalent SARS-CoV-2 variants, appearing greater for Delta than Omicron. Vaccinated CYP with prior SARS-CoV-2 infection had near-total protection against infection with the Delta variant, with re-infection close to null for at least 100 days; currently insufficient data are available to make similar comment regarding the Omicron variant. However, >90% of children in UK are currently thought to have had SARS-CoV-2 infection (modelled antibody prevalence from December 7, 2020 to February 4, 2022 of 82% in children aged 8-11 years and 94% in children aged 12-15 years; https://www.ons.gov.uk/peoplepopulationandcommunity/healthandsocialcare/conditionsanddiseases/bulletins/coronaviruscovid19infectionsurveyantibodyandvaccinationdatafortheuk/23february2022); thus our finding here may inform discussions regarding two-dose vaccination policy, particularly for CYP with prior infection.

Our community data regarding effectiveness of single-dose vaccination (at least for the considered time-scale) adds to previous publications. Phase 3 randomised trials in persons aged 12 years and older demonstrated high two-dose vaccine efficacy (95% CI, 75.3 to 100%^6^) with immunogenicity against earlier SARS-CoV-2 wild-type and Alpha variants^6, 7^ and protection against severe COVID-19 ^11, 20^. The PROTECT trial, which included 243 participants aged 12-17 years, estimated adjusted vaccine effectiveness of 94% (83%-98%) after two doses of BNT162b2^21^. Community data on vaccine effectiveness in adolescents are limited ^22, 23^. A national study in Israel observed that, after two weeks since administration of the second vaccine dose, effectiveness was 87.1%−91.2%. Until end of August 2021, none of the vaccinated and 33 of 9,969 unvaccinated adolescents who tested positive for SARS-CoV-2 (0.33%) were hospitalised for severe illness ^24^.

Our data suggesting a variant-dependent difference in vaccine efficacy concords with a recent pre-printed study in England ^25^, that used a test-negative case-control design to assess PCR-confirmed COVID-19. This study showed two-dose BNT162b2 vaccination in 12- to 17-year-old CYP was less effective against symptomatic disease from Omicron vs. Delta SARS-CoV-2 variants (variant genotype assumed from prevalence in population), with vaccine effectiveness against COVID-19 peaking 14 days after first dose for Delta and Omicron at 75-76%% and 50-51%% respectively, and decreasing to 36-48% and 23-27% by 8 weeks.

Our finding of a strong and sustained effect of prior SARS-CoV-2 infection on post-vaccination efficacy concurs with other studies also showing the effect of prior infection upon immunity. In one such, 32 participants with prior SARS-CoV-2 infection were assessed post-vaccination and evidenced strong neutralising antibody responses after first mRNA vaccination, that did not increase further following a second dose, and was higher than uninfected participants who had received two vaccine doses (n=139) ^26^.

Vaccination also resulted in a modest change in COVID-19 profile. Vaccinated CYP with post-vaccination SARS-CoV-2 infection appeared to have milder disease (i.e., fewer symptoms) than unvaccinated CYP, at least during the time of Delta predominance. Less difference was evident during Omicron predominance, especially in young people, noting here that disease appeared milder during Omicron predominance compared with Delta predominance, in both vaccinated and unvaccinated CYP. Relevantly, our data in unvaccinated CYP are consistent with our earlier study ^3^, and that of others (https://www.ons.gov.uk/peoplepopulationandcommunity/healthandsocialcare/conditio nsanddiseases/articles/coronaviruscovid19latestinsights/Overview#age), in that COVID-19 in children is usually milder, and of shorter duration, compared with adults ^27^, with much lower risk of severe disease, death and prolonged symptoms. Thus, *a priori* the capacity to show a large impact of vaccination on disease profile in CYP (in contrast to adults) was limited. Nonetheless, even during Omicron predominance odds ratios showed several symptoms were significantly less common in vaccinated vs. unvaccinated children. Our descriptive data of hospital presentations showed slightly lower numbers in vaccinated vs. unvaccinated children (0·64% vs. 1·58% for Omicron) but the opposite finding in young people (2·13% vs. 1·50% for Omicron). No robust comparisons can be made here.

Post-vaccination side-effects were common but dominated by local symptoms (70·8% of children, 60·2% of young people) which resolved rapidly in most cases. Our data here are very similar to those of the v-safe study (60%), a USA smartphone-based surveillance study of adolescents aged 12-17 years (N = 129,059) who completed at least one health check-in survey within one week after receiving a first dose of BNT162b2 vaccine ^10^. Systemic side-effects in our cohort were uncommon, non-critical, and non-persistent. The profile of systemic side-effects we observed was similar to that recorded in the pivotal clinical trial for the BNT162b2 vaccine ^6^ but much less common (e.g., for headache: 2·0%-3·7% in our cohort vs. 55·0-65·0% in the pivotal trial; for fatigue: 1·8%-3·3% vs. 60·0-66·0%). In the v-safe study, headaches were reported in 25-30%, and fatigue in 27-34% ^10^. It may be that only more severe systemic side effects were proxy-reported in our study. However, the nocebo effect may also be relevant in explaining the discrepancy (i.e., that individuals warned of potential side-effects may report them even when receiving the placebo), with a recent meta-analysis reporting extremely common nocebo responses in trials of COVID-19 vaccines (76.0% of systemic effects after first dose and 51.8% after second dose) ^28^.

The strengths of our study include community, real-time data generated from thousands of app users and a stringent study design. However, we also acknowledge the weaknesses of our study. Our data are generated by a volunteer citizen-science initiative delivered through an app, with the CSS cohort more likely to be of White Caucasian ethnicity and higher family socioeconomical status than the overall UK population ^14^. By virtue of UK government policy timing and age-tiered approach, our study was restricted to considering single-dose vaccination with BNT162b2 in CYP aged 12-17 years; our results may not apply to other vaccines or in younger age-groups. Our study was conducted with the prevailing SARS-CoV-2 variants Delta and Omicron; the impact of vaccination (both effectiveness and durability) and the profile of post-vaccination COVID-19 might differ with other variants. Indeed, the durability of vaccine protection against Omicron beyond 3 months in our cohort remains to be determined, due to insufficient data, noting here the ability to assess vaccination effectiveness in CYP naïve to SARS-CoV-2 is dwindling rapidly, given widespread infection in the community, particularly CYP. Vaccinated CYP were also much more likely to be tested for SARS-CoV-2 than unvaccinated CYP, for reasons that we are unable to determine. Our data do not allow determination of causal variants for CYP testing positive either post- or prior to vaccination: we can only infer likely variant based on population prevalences at time of testing. Vaccination can attenuate viral RNA shedding ^29^; this might result in the overestimation of vaccine effectiveness by reducing viral detection among infected individuals who received vaccination. In addition, divergence of infection risk soon after vaccination may indicate that the vaccinated had a lower risk of infection than the unvaccinated. Pertinently here, we could not incorporate into our analysis behaviours affecting risk of infection, such as mask-wearing, distancing, quarantines, and school attendance. Importantly, although myo- or peri-carditis have appeared as significant adverse effects in younger populations ^9^, no dedicated question assessed this specifically; however, our data regarding chest pain and dyspnoea were assessed and free text reports scrutinised for such adverse diagnoses.

Importantly, this study should be regarded as an interim report of the effects of one-dose BNT162b2 vaccine. Pivotal clinical trials ^6, 7^, product licensing, and most national recommendations indicate that more comprehensive immunization is achieved after at least two time-spaced dose administrations of BNT162b2 or other SARS-CoV-2 vaccinations. However, our data suggest some nuance may be appropriate regarding recommendations for second-dose vaccination, considered temporally (particularly with reference to prior infection) and to new variant emergence.

## Conclusions

One dose of BNT162b2 vaccine gives protection against SARS-CoV-2 infection for at least 90 days in CYP aged 12-17 years. Protection was lower for Omicron compared to Delta variants. Vaccine protection is modulated by the SARS-CoV-2 variant type, and by previously-acquired infection-induced immunity. Overall, BNT162b2 vaccine is well tolerated in children and young people, with common but fast-fading local effects, and uncommon and transient systemic effects. Severity of COVID-19 presentation after one dose of vaccine is generally milder, although unvaccinated CYP generally have an uncomplicated course, too. We contribute data for the benefit of governments and public health agencies, to be considered within the debate on vaccination in children, and for designing public health strategies to control future developments of the COVID-19 pandemic.

### Data sharing

Data collected in the COVID Symptom Study smartphone application may be shared with other health researchers through the UK National Health Service-funded Health Data Research UK (HDRUK) and Secure Anonymised Information Linkage consortium, housed in the UK Secure Research Platform (Swansea, UK).

Anonymised data are available to be shared with researchers according to their protocols in the public interest (https://web.www.healthdatagateway.org/dataset/fddcb382-3051-4394-8436-b92295f14259).

## Supporting information

STROBE checklist

## Data Availability

Data collected in the COVID Symptom Study smartphone application may be shared with other health researchers through the UK National Health Service-funded Health Data Research UK (HDRUK) and Secure Anonymised Information Linkage consortium, housed in the UK Secure Research Platform (Swansea, UK). Anonymised data are available to be shared with researchers according to their protocols in the public interest (https://web.www.healthdatagateway.org/dataset/fddcb382-3051-4394-8436-b92295f14259).

## Acknowledgments

This work is supported by the Wellcome Engineering and Physical Sciences Research Council (EPSRC) Centre for Medical Engineering at King’s College London (WT 203148/Z/16/Z) and the UK Department of Health via the National Institute for Health Research (NIHR) comprehensive Biomedical Research Centre (BRC) award to Guy’s & St Thomas’ NHS Foundation Trust in partnership with King’s College London and King’s College Hospital NHS Foundation Trust, the Medical Research Council (MRC) and British Heart Foundation. SO and MM are supported by the UK Research and Innovation London Medical Imaging & Artificial Intelligence Centre for Value Based Healthcare. SO is also supported by the Wellcome Flagship Programme (WT213038/Z/18/Z) and British Hearth Foundation. EM is funded by an MRC Skills Development Fellowship Scheme at KCL. Zoe Limited supported all aspects of building and running the app and service to all users worldwide.

## Author contributions

EM, LCa, KK performed the analyses

EM, LCa, CJS, SO, ELD worked at conceptualization and methodology LCa, JD performed data extraction and curation

EM, LCa ELD, MAb wrote the manuscript

BM, LCh, JCP, LP, AM, JW developed the data collection system SO, TDS, CJS conceived the CSS and obtained funds

EM, ELD coordinated this research

All the authors critically reviewed the manuscript.

All the authors had access to the COVID Symptom Study dataset, which is also accessible to researchers in the public interest.

EM, LCa, JD, MAn verified the data of this study.

## Declaration of interests

JCP, LP, AM, JW are employees of Zoe Limited.

TDS reports being a consultant for Zoe Limited, during the conduct of the study. All other authors have nothing to declare.

## Abbreviations

BNT162b2: Comirnaty SARS-CoV-2 vaccine (BioNTech, Pfizer)
CYP: Children and young people.
KCL: King’s College London
LFAT: Lateral flow antigen test
PCR: Polymerase chain reaction.
SARS-CoV-2: Severe acute respiratory syndrome-related coronavirus 2
UK: United Kingdom of Great Britain and Northern Ireland

## SUPPLEMENTARY DOCUMENT

**Supplementary Table 1.** List of symptom questions asked by the COVID Symptom Study application during the current study period.

| Symptom | COVID Symptom Study app question |
| --- | --- |
| Fever | Fever (at least 37.8C or 100F) |
| Persistent Cough | Persistent cough (coughing a lot for more than an hour or 3 or more coughing episodes in 24 hours) |
| Fatigue | Unusual fatigue (no; mild fatigue; severe fatigue/ I struggle to get out of bed) |
| Dyspnoea | Shortness of breath or trouble breathing (no; yes mild symptoms/ slight shortness of breath during ordinary activity: yes significant symptoms/ breathing is comfortable only at rest; yes, severe symptoms/ breathing is difficult even at rest). |
| Anosmia/Ageusia | Loss of smell / taste |
| Hoarse Voice | Unusually hoarse voice |
| Chest Pain | Unusual chest pain or tightness in your chest |
| Abdominal Pain | Unusual abdominal pain or stomach-ache |
| Diarrhoea | Diarrhoea |
| Delirium | Confusion, disorientation or drowsiness |
| Eye Soreness | Do your eyes have any unusual eye-soreness or discomfort (e.g. light sensitivity, excessive tears, or pink/red eye)? |
| Low appetite (anorexia) | Skipping meals |
| Headache | Headache |
| Nausea | Nausea or vomiting |
| Dizziness | Dizziness or light-headedness |
| Sore Throat | Sore or painful throat |
| Myalgias | Unusual strong muscle pains or aches |
| Red Welts | Raised, red, itchy welts on the skin or sudden swelling of the face or lips |
| Blisters | Red/purple sores or blisters on your feet, including your toes |
| Rashes | Rash on your arms or torso |
| Sensitive Skin | Strange, unpleasant sensations in your skin like pins & needles or burning |
| Hair Loss | Unusual hair loss |
| Low mood (depression) | Feeling down, depressed, or hopeless |
| Brain Fog | Loss of concentration or memory (brain fog) |
| Dysosmia/Dysgeusia | Altered smell / taste (things smell or taste different to usual) |
| Rhinorrhoea | Runny nose |
| Sneezing | Sneezing more than usual |
| Ear Pain | Earache |
| Tinnitus | Ringling in your ears |
| Lymphadenopathy | Swollen neck glands |
| Palpitations | Unusually fast or irregular heartbeat (palpitations) |
| Arthralgias | Unusual joint pains or aches |
| Mouth Ulcers | Mouth or tongue ulcers |
| Tongue Changes | Changes to tongue surface |

**Supplementary Table 2.** List of questions about local symptoms at the site of vaccination, asked by the COVID Symptom Study application during the current study period.

| **Symptom** | **Question asked through the app:** Are you experiencing any symptoms near the injection site? (Check all that apply). |
| --- | --- |
| **Pain** | Pain |
| **Redness** | Redness |
| **Swelling** | Swelling |
| **Axillary lymphadenopathy** | Swollen glands in the armpit |
| **Warmth** | Warmth |
| **Pruritis** | Itch |
| **Tenderness** | Tenderness |
| **Ecchymoses** | Bruising |
| **Other** | Other: (free text) |

**Supplementary Table 3.** Cohort selection criteria for each analysis and associated sample sizes.

| Selection criteria | 12-15-year-old group (children) | 16-17-year-old group (young people) |
| --- | --- | --- |
| **Post-vaccination infection risk.**<br>**Vaccinated:**<br>- One dose of BNT162b2 vaccination.<br>- Absence of documented infection prior to vaccination.<br>- SARS-CoV-2 test at least 14 days after one dose of BNT162b2 vaccination.<br>- First SARS-CoV-2 positive test (if any), and any negative test before the first positive.<br>**Unvaccinated:**<br>- First SARS-CoV-2 positive test (if any), and any negative test before the first positive. | Delta=5,658<br>Omicron=6,416<br>Delta=3,977<br>Omicron=723 | Delta=4,857<br>Omicron=857<br>Delta=816<br>Omicron=207 |
| **Infection over time:**<br>**- Vaccinated CYP without prior SARS-CoV-2 infection:**<br>- One dose of BNT162b2 vaccination.<br>- Absence of documented infection prior to vaccination.<br>- SARS-CoV-2 test any time after one dose of BNT162b2 vaccination.<br>- First SARS-CoV-2 positive test (if any), and any negative test before the first positive.<br>**- Unvaccinated (with tests matched to vaccinated CYP without prior infection 1:1 by week of test):**<br>- First SARS-CoV-2 positive test (if any), and any negative test before the first positive.<br>**- Vaccinated CYP with prior SARS-CoV-2 infection matched to vaccinated CYP without prior SARS-CoV-2 infection 1:1 by week of test):**<br>- One dose of BNT162b2 vaccination.<br>- Documented positive test result prior to vaccination.<br>- Further positive SARS-CoV-2 test any time after one dose of BNT162b2 vaccination.<br>- First SARS-CoV-2 positive test (if any), and any negative test before the first positive.<br>**- Vaccinated without prior infection (with tests** | Delta=4,240<br>Omicron=1,339<br>Delta=11,156<br>Omicron=1,339<br>Delta=479<br>Omicron=717 | Delta=9,048<br>Omicron=262<br>Delta=2,132<br>Omicron=262<br>Delta=963<br>Omicron=22 |
| ***matched to vaccinated previously infected CYP, matched 1:1 by week of test, then age and gender):***<br>- First SARS-CoV-2 positive test (if any), and any negative test before the first positive. | Delta=479<br>Omicron=710 | Delta=963<br>Omicron=29 |
| **Post-vaccination infection profile (including hospital presentation)**<br>Populations matched* by age, gender week of test and BMI.<br>**Vaccinated:**<br>- One dose of BNT162b2 vaccination.<br>- Absence of documented infection prior to vaccination.<br>- SARS-CoV-2 test at least 14 days after one dose of BNT162b2 vaccination.<br>- First SARS-CoV-2 positive test.<br>**Unvaccinated:**<br>- First SARS-CoV-2 positive test<br>* Matching performed using Euclidean distance based on the variables mentioned above. 1:1 matched population not performed. | Delta=320<br>Omicron=1,269<br>Delta=320<br>Omicron=409 | Delta=564<br>Omicron=166<br>Delta=564<br>Omicron=97 |
| **Local and systemic post-vaccination symptoms**<br>- One dose of BNT162b2 vaccination.<br>- Absence of documented infection prior to vaccination.<br>- At least one report of symptoms (including their absence) within 7 days after vaccine uptake. | Systemic Symptoms = 7,809<br>Local Symptoms = 7,751 | Systemic Symptoms= 3,189<br>Local Symptoms = 2,876 |

**Supplementary Table 4.** Odds ratios for symptom prevalence in vaccinated vs. unvaccinated CYP, first testing positive for SARS-CoV-2 during periods of Delta (left) and Omicron (right) variant predominance. Asymptomatic CYP testing positive for SARS-CoV-2 are included in the computation. Bold encodes significant p-values after false discovery rate correction (alpha =0.05).

|  | CYP: 12-15 yo |  |  |  | CYP: 16-17 yo |  |  |  |
| --- | --- | --- | --- | --- | --- | --- | --- | --- |
|  | Delta |  | Omicron |  | Delta |  | Omicron |  |
| Symptoms | OR [IQR] | p-value | OR [IQR] | p-value | OR [IQR] | p-value | OR[IQR] | p-value |
| Abdominal pain | 0.71 [0.43; 1.17] | 0.1770 | **0.49 [0.35; 0.70]** | **0.0001** | **0.50 [0.33; 0.77]** | **0.0015** | 0.57 [0.21; 1.57] | 0.2791 |
| Dysosmia | 0.67 [0.44; 1.00] | 0.0501 | **0.59 [0.41; 0.85]** | **0.0044** | 0.82 [0.62; 1.09] | 0.1637 | 1.31 [0.50; 3.43] | 0.5878 |
| Blisters on feet | 0.33 [0.03; 3.20] | 0.3371 | 0.62 [0.11; 3.43] | 0.5830 | 0.24 [0.03; 2.01] | 0.1876 | 0 [0;inf] | 0.9993 |
| Brain Fog | 0.76 [0.40; 1.43] | 0.3937 | 0.59 [0.38; 0.91] | 0.0175 | 0.76 [0.53; 1.10] | 0.1454 | 0.73 [0.26; 2.10] | 0.5622 |
| Chest Pain | 0.72 [0.36; 1.43] | 0.3489 | 0.77 [0.48; 1.24] | 0.2867 | **0.54 [0.36; 0.82]** | **0.0040** | 0.65 [0.21; 2.08] | 0.4718 |
| Chills or Shivers | 0.63 [0.42; 0.93] | 0.0217 | 0.75 [0.56; 1.00] | 0.0481 | 0.75 [0.56; 0.99] | 0.0455 | 0.68 [0.34; 1.33] | 0.2573 |
| Confusion | **0.22 [0.07; 0.67]** | **0.0073** | **0.46 [0.26; 0.82]** | **0.0079** | 0.67 [0.43; 1.04] | 0.0724 | 0.44 [0.14; 1.33] | 0.1453 |
| Diarrhoea | 0.44 [0.22; 0.89] | 0.0231 | 0.81 [0.47; 1.39] | 0.4451 | **0.55 [0.34; 0.88]** | **0.0128** | 0.98 [0.36; 2.73] | 0.9749 |
| Dizziness | **0.48 [0.31; 0.73]** | **0.0008** | **0.52 [0.39; 0.70]** | **<0.0001** | **0.59 [0.43; 0.81]** | **0.0010** | 0.43 [0.22; 0.86] | 0.0173 |
| Tinnitus | 0.72 [0.24; 2.11] | 0.5481 | 0.77 [0.44; 1.36] | 0.3736 | 0.60 [0.33; 1.08] | 0.0896 | 0.47[0.13; 1.72] | 0.2519 |
| Earache | 0.48 [0.23; 1.01] | 0.0536 | 0.60 [0.38; 0.95] | 0.0305 | 0.70 [0.44; 1.10] | 0.1240 | 0.33 [0.11; 0.99] | 0.0477 |
| Eyes soreness | **0.39 [0.24; 0.64]** | **0.0002** | **0.62 [0.43; 0.88]** | **0.0082** | **0.49 [0.36; 0.69]** | **<0.0001** | 0.73 [0.31; 1.69] | 0.4559 |
| Fatigue | **0.66 [0.48; 0.90]** | **0.0097** | 0.83 [0.65; 1.03] | 0.0868 | **0.64 [0.50; 0.82]** | **0.0003** | 0.66 [0.39; 1.12] | 0.1221 |
| Low Mood | 0.64 [0.33; 1.24] | 0.1832 | 0.65 [0.39; 1.08] | 0.0962 | **0.61 [0.40; 0.94]** | **0.0230** | 1.03 [0.32; 3.35] | 0.9611 |
| Fever | 0.71 [0.49; 1.02] | 0.0642 | **0.67 [0.52; 0.87]** | **0.0029** | **0.56 [0.42; 0.73]** | **<0.0001** | 0.78 [0.41; 1.50] | 0.4623 |
| Hair Loss | 0.95 [0.06;<br>15.66] | 0.9699 | 1.24 [0.13;<br>11.59] | 0.8492 | 0.78 [0.28; 2.18] | 0.6394 | **1 [1;1]** | **0** |
| Headache | **0.55 [0.40; 0.75]** | **0.0002** | **0.76 [0.60; 0.95]** | **0.0148** | **0.61 [0.48; 0.78]** | **0.0001** | 0.76 [0.45; 1.29] | 0.3169 |
| Hoarse voice | 0.74 [0.45; 1.20] | 0.2206 | 1.12 [0.79; 1.60] | 0.5343 | **0.61 [0.44; 0.84]** | **0.0026** | 1.14 [0.48; 2.70] | 0.7671 |
| Palpitations | 0.72 [0.23; 2.29] | 0.5758 | 1.09 [0.46; 2.57] | 0.8459 | 0.74 [0.37; 1.47] | 0.3844 | 2.08 [0.22;<br>19.74] | 0.5246 |
| Anosmia | 0.79 [0.54; 1.16] | 0.2213 | **0.48 [0.31; 0.73]** | **0.0007** | 0.81 [0.62; 1.05] | 0.1138 | 0.53 [0.20; 1.36] | 0.1852 |
| Nausea | **0.35 [0.20; 0.59]** | **0.0001** | 0.72 [0.50; 1.02] | 0.0652 | **0.51 [0.35; 0.74]** | **0.0004** | 0.54 [0.23; 1.27] | 0.1588 |
| Persistent Cough | 0.74 [0.52; 1.05] | 0.0873 | 1.19 [0.90; 1.59] | 0.2264 | **0.73 [0.57; 0.94]** | **0.0140** | 1.79 [0.87; 3.67] | 0.1132 |
| Rashes | 0.36 [0.10; 1.39] | 0.1394 | 0.66 [0.29; 1.51] | 0.3296 | **0.27 [0.11; 0.72]** | **0.0080** | 0 [0; inf] | 0.9992 |
| Red Welts FL | 1.00 [0.14; 7.14] | 0.9989 | 0.64 [0.26; 1.62] | 0.3507 | 0.43 [0.19; 0.98] | 0.0443 | 1.10 [0.16; 7.58] | 0.9226 |
| Rhinorrhoea | **1.54 [1.13; 2.11]** | **0.0069** | 0.96 [0.76; 1.20] | 0.7035 | 1.29 [1.02; 1.64] | 0.0347 | 0.92 [0.53; 1.60] | 0.7692 |
| Dyspnoea | 0.81 [0.43; 1.52] | 0.5043 | 0.97 [0.59; 1.58] | 0.8868 | **0.65 [0.44; 0.96]** | **0.0289** | 1.21 [0.42; 3.46] | 0.7287 |
| Sensitive Skin | 0.53 [0.19; 1.46] | 0.2215 | 0.68 [0.35; 1.30] | 0.2439 | **0.43[0.24; 0.77]** | **0.0043** | 0.13 [0.03; 0.67] | 0.0143 |
| Low Appetite | **0.47 [0.29; 0.76]** | **0.0018** | **0.54 [0.39; 0.75]** | **0.0002** | **0.40 [0.28; 0.57]** | **0.0000** | 0.71 [0.33; 1.54] | 0.3855 |
| Sneezing | 1.12 [0.79; 1.59] | 0.5174 | 0.91 [0.70; 1.18] | 0.4751 | **1.28 [1.00; 1.67]** | **0.0609** | 1.26 [0.68; 2.34] | 0.4551 |
| Sore throat | 0.85 [0.62; 1.17] | 0.3161 | 0.92 [0.74; 1.16] | 0.4899 | **0.62 [0.49; 0.79]** | **0.0001** | 0.77 [0.45; 1.29] | 0.3152 |
| Lymphadenopathy | 0.65 [0.34; 1.24] | 0.1901 | 0.77 [0.48; 1.24] | 0.2849 | **0.38 [0.24; 0.61]** | **0.0001** | 1.01 [0.39; 2.61] | 0.9821 |
| Myalgias | **0.49 [0.29; 0.84]** | **0.0099** | **0.48 [0.34; 0.68]** | < 0.0001 | **0.50 [0.35; 0.73]** | **0.0003** | 0.56 [0.27; 1.18] | 0.1282 |
| Arthralgias | **0.33 [0.19; 0.56]** | **0.0000** | 0.72 [0.52; 1.00] | 0.0474 | **0.58 [0.42; 0.80]** | **0.0010** | 0.39 [0.20; 0.76] | 0.0057 |

**Supplementary Figure 1.**
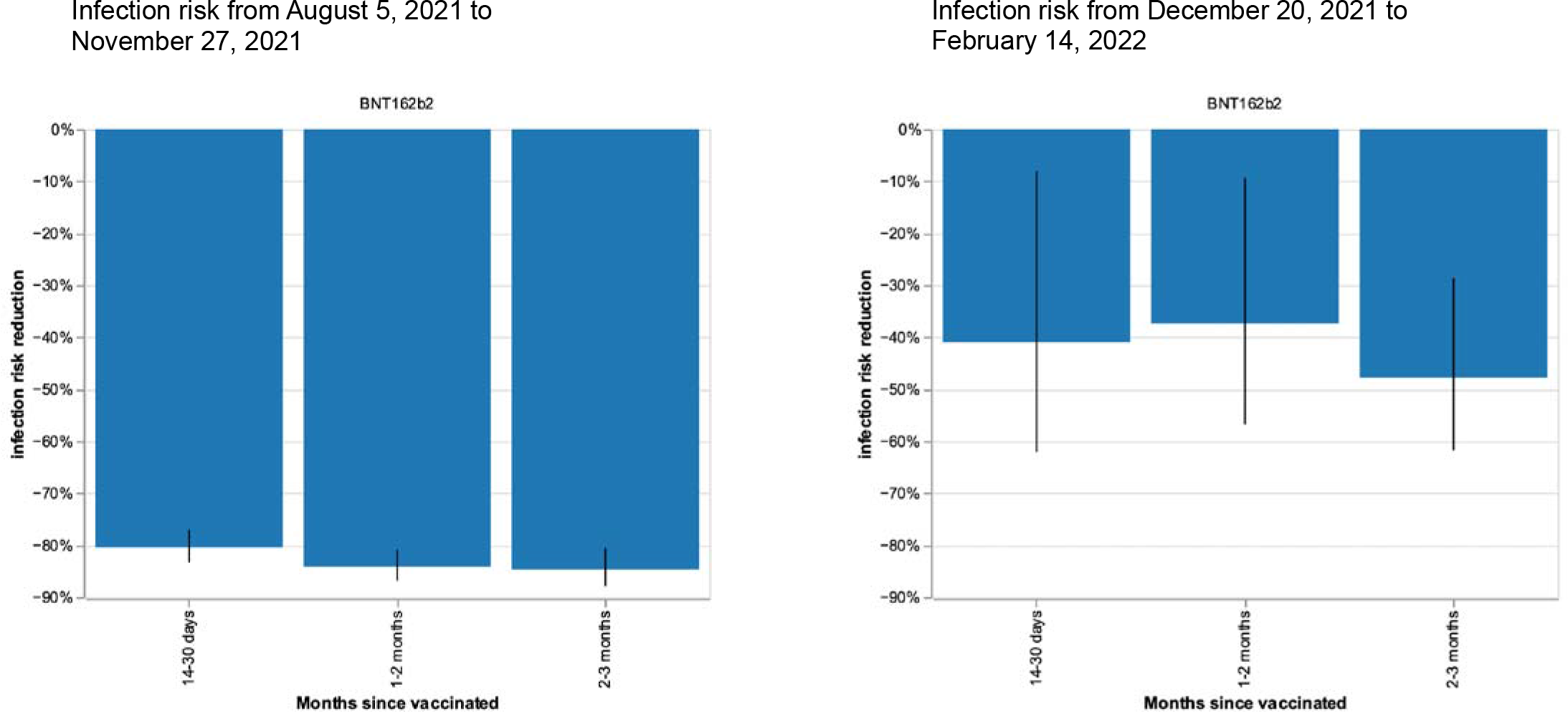
Infection risk reduction after first dose BNT162b2 vaccination in young people aged 16-17 years, during periods of Delta (left) and Omicron (right) variant predominance in UK. Bars represent risk reduction at 14-30 days, 1-2 months, and 2-3 months in young people post-vaccination, compared with unvaccinated young people. Black lines represent 95% CIs. Number of observations (i.e., tests) of young people: during Delta variant predominance in UK: n(test) = 5,673 (816 unvaccinated, 4,857 vaccinated); during Omicron variant predominance: n(test) = 1,064 (207 unvaccinated, 857 vaccinated).

**Supplementary Figure 2.**
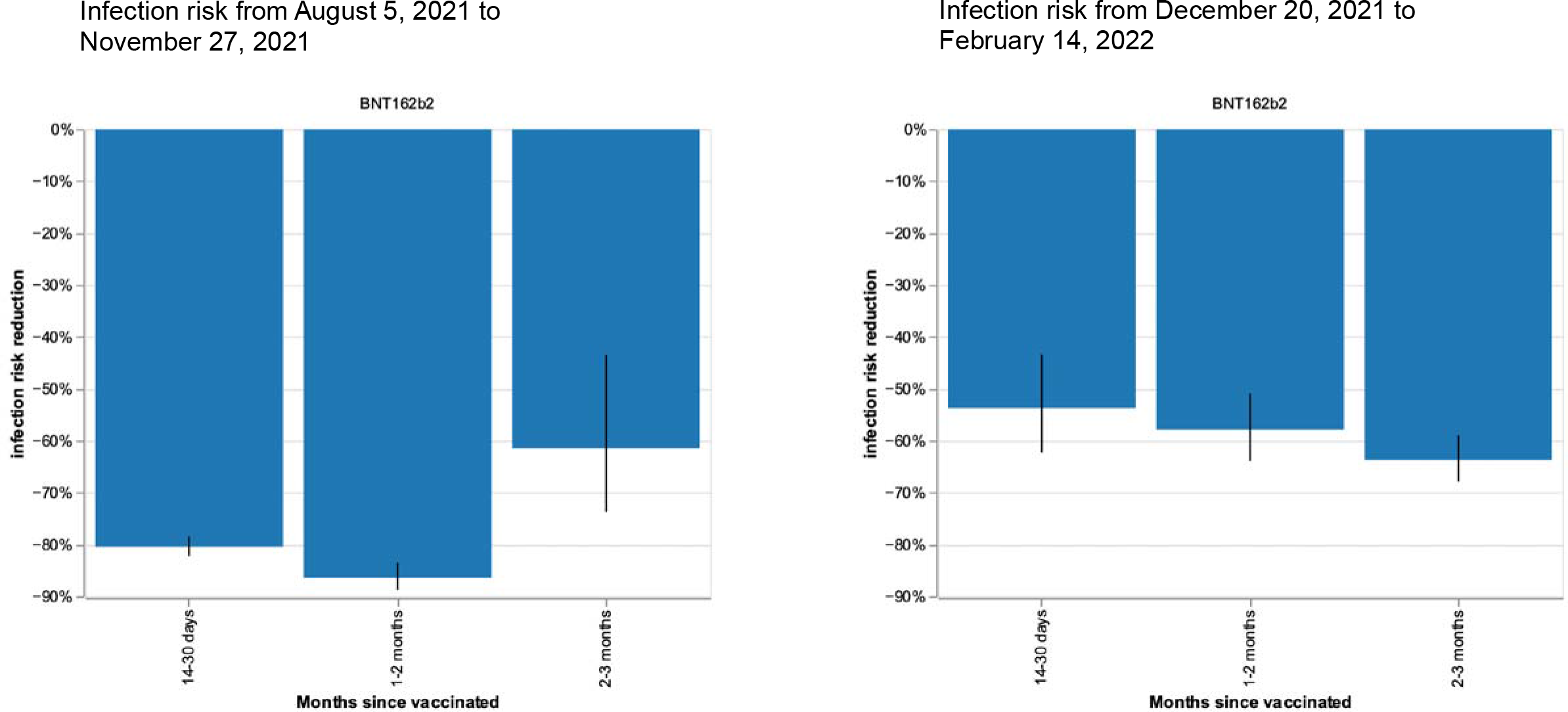
Infection risk reduction after first dose BNT162b2 vaccination in children aged 12-15 years, during periods of Delta (left) and Omicron (right) variant predominance in UK. Bars represent risk reduction at 14-30 days, 1-2 months, and 2-3 months for post-vaccination infection, compared with unvaccinated children. The black lines show 95% CIs. Number of observations (i.e., tests) of children: for Delta variant predominance, n(test) = 9,635 (3,977 unvaccinated, 5,658 vaccinated); for Omicron variant predominance, n(test) = 7,139 (723 unvaccinated, 6,416 vaccinated).

**Supplementary Figure 3.**
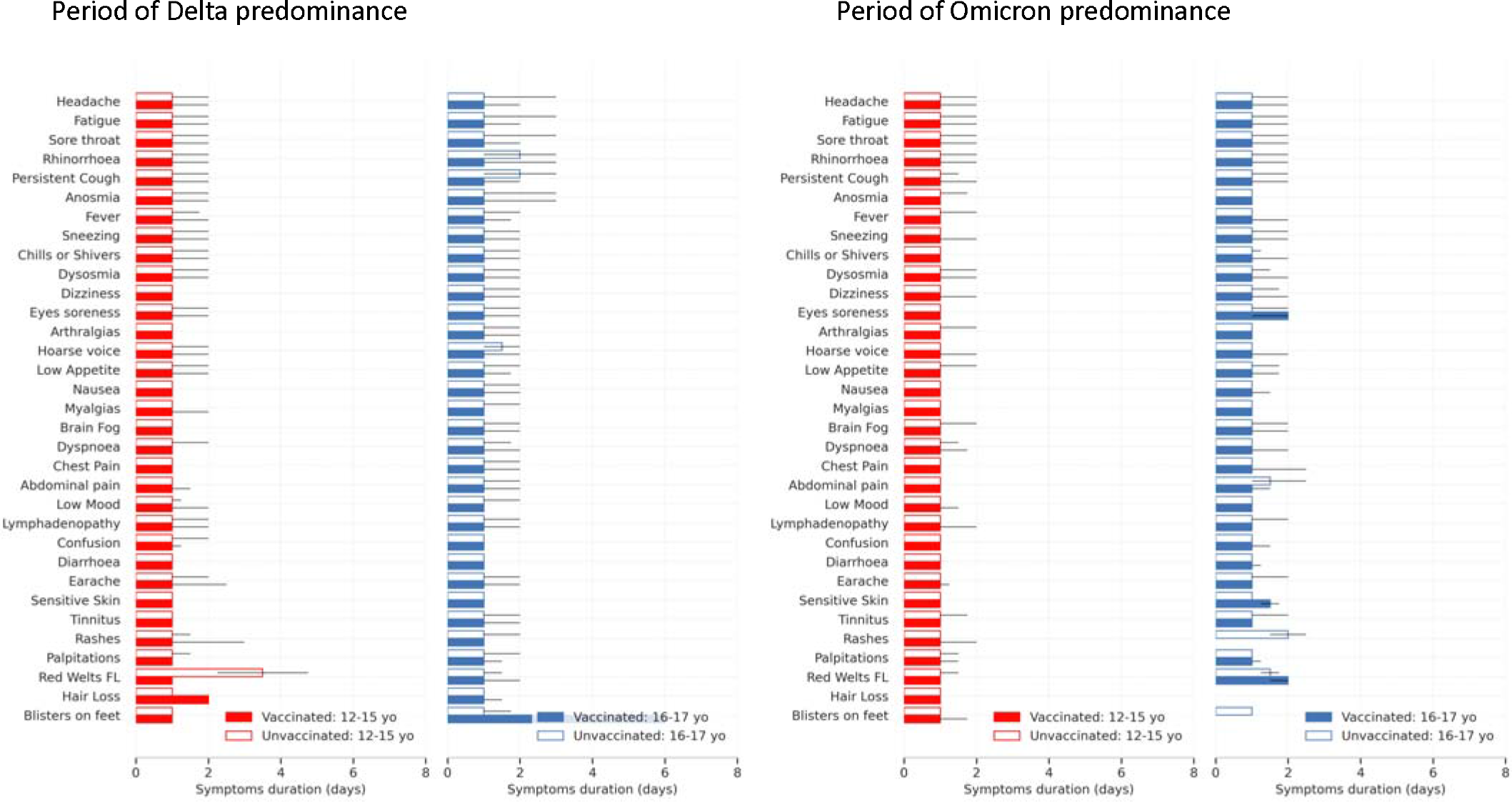
Symptom duration in vaccinated and unvaccinated CYP with symptomatic *de novo* SARS-CoV-2 infection during periods of Delta (left panels) and Omicron (right panels) variant predominance. Asymptomatic CYP testing positive for SARS-CoV-2 are excluded from computation of symptom duration. Bars show median duration; black lines show IQR.

**Supplementary Figure 4.**
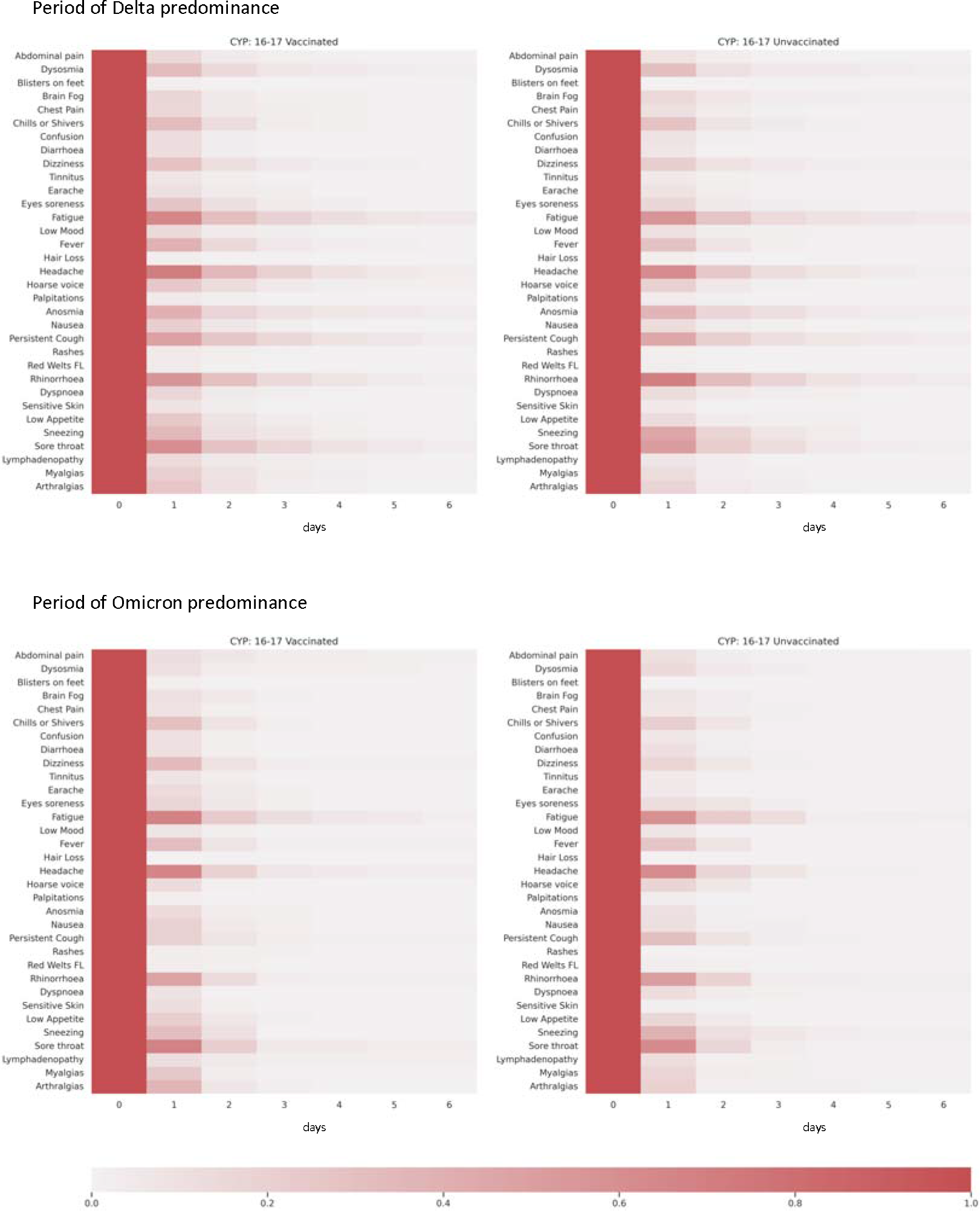
Persistence of individual symptoms over 7 days after testing positive for *de novo* SARS-CoV-2, in vaccinated (left) and unvaccinated (right) young people aged 16- 17 years, during periods of Delta (top) and Omicron (bottom) variant prevalence in UK. Each row represents persistence over time (in days) of each symptom for individuals reporting that symptom. Day 0 is defined as the first day of symptom presentation.

**Supplementary Figure 5.**
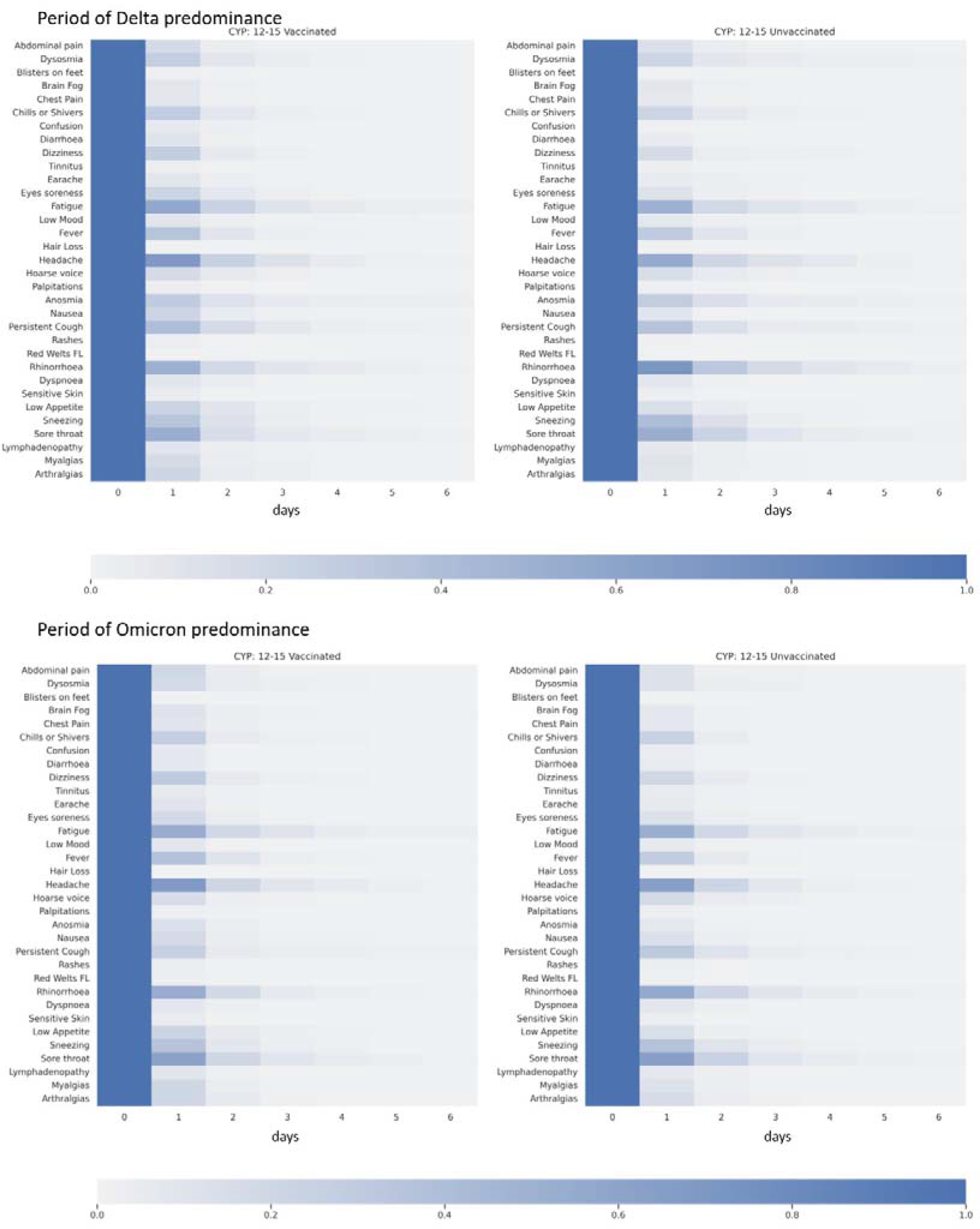
Persistence of individual symptoms over 7 days after testing positive for *de novo* SARS-CoV-2, in vaccinated (left) and unvaccinated (right) children aged 12-15 years old, during periods of Delta (top) and Omicron (bottom) variant prevalence in UK. Each row represents persistence over time (in days) of each symptom for individuals reporting that symptom. Day 0 is defined as the first day of symptom presentation.

**Supplementary Figure 6.**
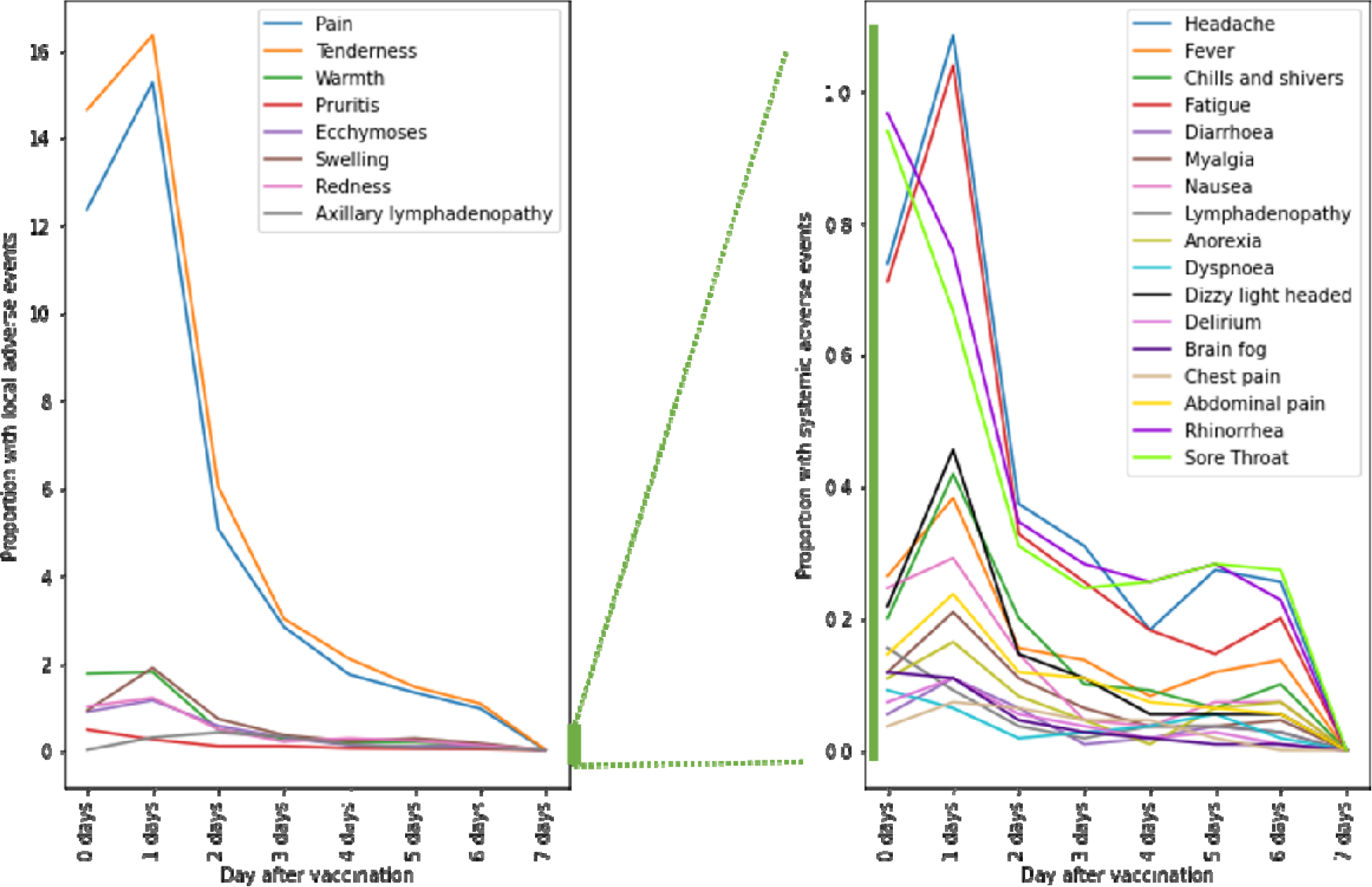
Proportion of CYP aged 12 to 17 years reporting vaccine side-effects after one dose BNT162b2. Local effects at the arm of injection considered for 7 days are presented in the left panel. Systemic symptoms presenting within the same 7 days after vaccination are presented in the right panel.

## Notes

### Author Declarations

Ethics approval was granted by the KCL Ethics Committee (LRS-19/20-18210).

